# Effectiveness of interactive dashboards to optimise prescribing in general practice: A systematic review

**DOI:** 10.1101/2024.08.22.24312420

**Authors:** Caroline McCarthy, Patrick Moynagh, Áine Mannion, Ashely Wei, Barbara Clyne, Frank Moriarty

## Abstract

**Background:** The World Health Organisation’s Medication Without Harm campaign aims to reduce severe avoidable medication related harm by 50%. This systematic review explored the characteristics of interventions that provide visual and longitudinal feedback on prescribing, or interactive dashboards, in general practice and the effect of these interventions on prescribing-related outcome measures.

**Methods:** This systematic review was registered prospectively and reported in line with PRISMA guidelines. Multiple databases and grey literature were searched in November 2023 to identify interventional studies that explored the effect of interactive dashboards on prescribing-related outcomes in general practice. Two independent researchers conducted screening, data extraction, and risk of bias assessment. Interventions were described narratively, and a random-effects meta-analysis was performed for comparable studies.

**Results:** Eight randomised controlled trials, one controlled before-and-after study and three interrupted time series were included. Six studies reported a significant positive effect on prescribing-related outcomes, with an effect seen more often for studies focusing on potentially inappropriate prescribing (PIP) (four out of six). Two of the six studies that focused on antibiotic prescribing demonstrated a significant effect. A meta-analysis of three RCTs involving 160 general practices and 198,135 patients demonstrated the overall odds of PIP was 0.91 (95%CI: 0.77-1.06 I^2^=71.8%) in the intervention compared to control group.

**Conclusion:** Interactive dashboards show promise for supporting safe and effective prescribing in general practice. Future research should focus on developing core outcome sets to facilitate future meta-analyses of effectiveness as well as optimising their implementation and understanding how to sustain user engagement.

**Lay summary:** The World Health Organization’s “Medication Without Harm” campaign aims to reduce preventable medication-related harm by 50%. One way to support safe prescribing is by giving prescribers ongoing feedback on their prescribing habits using interactive dashboards. These dashboards provide visual and long-term data to help guide safer and more effective prescribing. This research looked at how interactive dashboards have been used in general practice and whether their use improves prescribing. Researchers systematically searched the published literature and identified 12 relevant studies. Some studies involved randomly assigning doctors or practices to either use the dashboards or continue usual care. Others compared prescribing practices before and after introducing dashboards or looked at practices that used dashboards compared to those that did not. Six of the studies showed improvements in prescribing, especially when focused on reducing high-risk prescriptions. The data for three studies that looked at high-risk prescribing involving 160 general practices and 198,135 patients showed that interactive dashboards may reduce the chance of unsafe prescribing by 8.8%. However, this result was not statistically significant, and the difference in results between studies means the true effect remains uncertain. The findings highlight the potential of interactive dashboards to support safer prescribing in general practice, though further research is needed.

## Introduction

Prescribing is the most common healthcare intervention and advances in therapeutics have improved the lives and life expectancy of many people living with chronic illness.^1^ However, alongside these improvements there has been an increase in potentially inappropriate prescribing (PIP) which has potential negative consequences for individuals, society and healthcare systems.^2^ Prescribing for older patients with multiple chronic illnesses is particularly challenging as prescribers must contend with both potential drug-drug and drug-disease interactions.^3^ The reasons for PIP are complex and multifaceted and include systems failures, particularly at the interface between primary and secondary care, individual patient factors such as increasing age and multimorbidity, clinician factors such as knowledge and attitudes and broader societal attitudes about the roles and benefits of medicines.^4^ To both measure and address this, a variety of explicit criteria have been developed for use in both research and clinical practice.^5^ These measures tend to focus on highly prevalent PIP or those with the potential for significant harm. More recently there has been a shift towards identifying and addressing low-value prescribing, which on a population level can result in harm both directly (e.g. experiencing adverse effects from medicines that are not providing benefit) and indirectly (e.g. contributing to non-adherence and through the opportunity cost of other cost-effective interventions not resourced due to spending on low-value medicines).^6^ The application of explicit measures of medication appropriateness have been demonstrated to be effective at improving prescribing and have the advantage of being relatively reproducible, reliable and easy to apply to large numbers of people.^7^

With recent advancements in electronic healthcare records (EHR) and prescribing, it is now possible to apply explicit criteria to routine prescribing or dispensing datasets. OpenPrescribing.net is a prominent example, where the vast amount of anonymous prescription data published by NHS England each month is analysed and presented on a web platform to allow for comparative benchmarking between practices.^8^ However, when using publicly available anonymous data, only a subset of explicit criteria can be applied as patient-level data such as age and co-morbidities are not included. In addition, it is challenging for clinicians to identify and act on individual instances of sub-optimal prescribing. One approach to combine both audit and feedback and clinical decision support (both of which have been identified as effective methods to improve prescribing^9,10^), while maintaining anonymity, has been to embed code within practice systems and export aggregate-level data.^11^ This aggregated data can then be fed back to individual practices in the form of interactive dashboards and allow for comparative benchmarking. This approach ensures that individual patient identities are protected while allowing practices to compare their performance, identify areas for improvement and act on individual instances of high-risk prescribing.

Given advancements in data infrastructure in general practice and the need to address both high-risk and low value prescribing, this systematic review aimed to explore characteristics and effectiveness of interactive dashboard interventions on prescribing outcomes in general practice with the additional goal of informing future intervention development and e-prescribing infrastructure.

## Methods

The methods have been described previously in our published protocol.^12^ This systematic review was prospectively registered on PROSPERO (CRD42023481475), conducted in line with guidance set out in the Cochrane Handbook for Systematic Reviews of Interventions,^13^ and reported in adherence to PRISMA statement, S1 Appendix.^14^

### Data sources and search strategy

A systematic literature search was conducted 22^nd^ November 2023 in the following databases; PubMed, EMBASE, MEDLINE (OVID), PsycINFO (EBSCOhost), CINAHL (EBSCOhost), Scopus and the Cochrane Library (OVID). This was supplemented by grey literature searches in OpenGrey, CADTH Grey Matters and the International Clinical Trials Registry Platform (ICTRP) as well as backward and forward citation chasing using an automated citation chaser.^15^ There were no restrictions placed on language or year of publication. Search terms included keywords to capture the intervention (e.g. “interactive dashboard”, “clinical audit”, “medical audit”, “benchmarking”, “data visualisation”) the population (e.g. “general practitioner”, “primary care*”) and the outcomes (e.g. “PIP”, “prescribing”). See S2 Appendix for electronic search reports.

### Eligibility criteria

All interventional designs were eligible for inclusion including randomised controlled trials (RCTs) (e.g. cluster RCTs, step wedged RCTs and individually randomised RCTs) and non-randomised interventional studies for example interrupted time series (ITS) design and controlled before and after studies as recommended by the Cochrane Effective Practice and Organisation of Care (EPOC) group, see S3 Appendix for a summary of inclusion and exclusion criteria. The population of interest was primary care prescribers including non-medical prescribers working in primary care (e.g. pharmacists). An interactive dashboard was defined as a platform designed to provide ongoing feedback of real-time (defined as no older than one year) prescribing data in a visual format and that allowed for comparative benchmarking against peers or a set standard. A true interactive dashboard allows direct manipulation of data with visual analytic tools, however studies that did not have an interactive element but provided feedback of multiple parameters and/or configurations from the dataset were also included. Simple clinical decision support interventions or audit and feedback interventions that did not give longitudinal and ongoing feedback of real-time data were both excluded. Multi-faceted interventions that included interactive dashboards alongside other components such as education, clinical decision support or targeted behavioural change strategies were included. The outcome of interest was any prescribing related outcome measure such as explicit prescribing criteria or prescribing rates where a higher rate is described as reflecting lower quality (e.g. antibiotic, benzodiazepine or opioid use).

### Study selection and data extraction

Identified records were uploaded to Covidence and independently assessed for inclusion based on title and abstract and then full text papers by two researchers (CMC, PM, AM, AW, FM), blinded to each other’s decisions, with disagreement resolved by consensus. Data was extracted independently by two researchers using a purposely developed data extraction tool in Covidence (CMC, PM), see S3 Appendix for a list of all data points extracted. Methodological quality assessment was assessed independently by two reviewers using the Cochrane EPOC risk of bias tool (CMC, PM).^16^

### Analysis

The Template for Intervention Description and Replication (TIDieR) checklist^17^ was used as a framework to narratively summarise interventions. For multi-faceted interventions this framework was utilised to describe the interactive dashboard component alone. We categorised effectiveness based on intervention type (for example if there was a truly interactive component to the dashboard or whether the dashboard was part of a multi-faceted intervention), outcome of interest (e.g. high-risk prescribing or antibiotic prescribing rates) and study design. A meta-analysis using a random-effects model was performed where at least two studies were comparable in terms of participants, study design and outcomes. If the desired direction of effect differed across outcomes, the measure of association (e.g., odds ratio) was inverted to ensure comparability. Heterogeneity across studies was assessed using the I² statistic, with an I² value greater than 50% considered indicative of substantial heterogeneity. Although 12 studies were identified, a funnel plot was not performed due to the heterogeneity in study design and outcomes. Instead, a narrative assessment was conducted, acknowledging the potential limitations in detecting publication bias.

## Results

### Search Results

A total of 12,918 records were identified from database searching and a further 197 from other sources. Following deduplication, 10,735 records were screened, with 121 full texts assessed for eligibility, and 12 studies, reported in 11 different papers, were included in the review, Figure 1. See S4 Appendix for a table of the excluded studies from full text review and their reason for exclusion.

**Figure 1.**
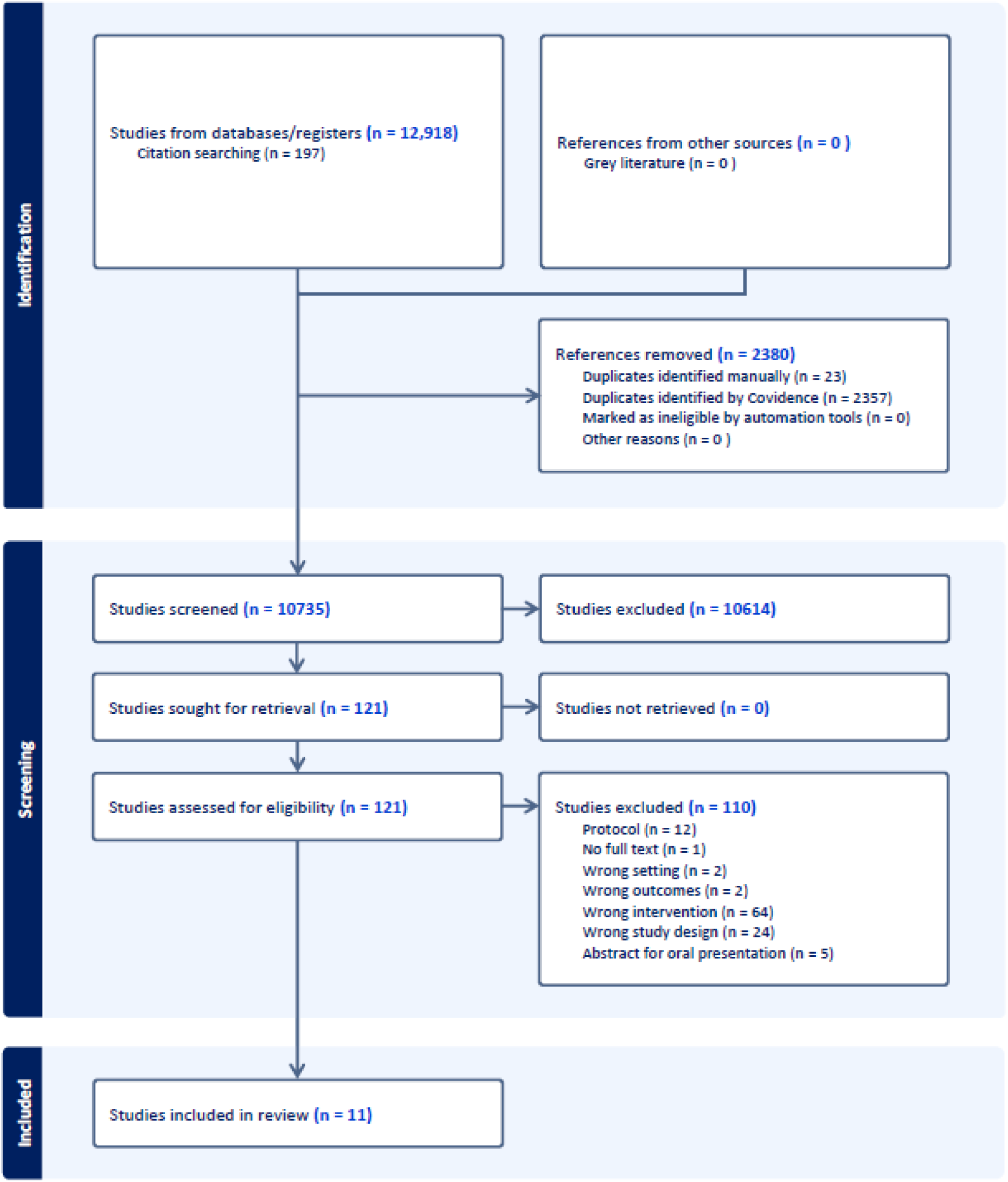
Study flow diagram. Characteristics of included studies

Of the 12 included studies, three were ITSs,^18–20^ one was a controlled before and after study^21^ and eight were RCTs,^22–28^ S6 Appendix Characteristics of Included Studies. Four of the RCTs were a cluster design.^23,25,27,28^ Two RCTs that targeted individual physician antibiotic prescribing were individually randomised at the physician level^22,26^ and the final RCT was a stepped wedge design where six clusters were sequentially allocated to the intervention every month.^24^ One paper described randomly assigning practices to one of two parallel cRCTs.^28^ In each cRCT, the control group in one trial served as the intervention group in the other, leading to a total of four distinct studies reported in the same paper. These studies were conducted simultaneously with the same intervention but focused on different outcomes. However, only two of these four studies were considered relevant for this systematic review, as the outcomes of the other two were not relevant. Therefore, both of the relevant studies from this paper were included in this review.^28^ In seven of the included studies, one ITS and six RCTs, participants (either individual prescribers or practices) were not aware of their participation in the study, as the interventions were implemented at a system level or as part of routine policy changes.^19,22,23,25–28^ In two additional studies it is unclear whether practices/physicians were recruited and consented^18,24^ and in the final two studies practices from a defined region were invited to participate.^20,21^ The number of participants included in the individual studies ranged from 12 to 1,401 practices and 43 to 3,426 physicians. None of the included studies recruited individual patients.

Six studies focused on antibiotic prescribing and used aggregated data, for example the rate of antibiotic prescriptions per 100 consultations or the proportion of all antibiotics that were broad-spectrum.^18,20,22–24,26^ The remaining six studies focused on PIP.^19,21,25,27,28^ Five of these reported the number of patients potentially at risk and affected. The remaining study reported the mean number of patients per practice with an inappropriate bronchodilator prescription, but did not provide the numerator and denominator.^27^

### Characteristics of interventions

The characteristics of the interactive dashboards of the included studies are described based on the TIDieR checklist in Table 1.^17^ The tailoring and modifications components of this framework were generally not applicable as these are often more relevant to implementation programmes, where interventions may need to be adapted to fit specific contexts or populations.^17^ The interventions identified in this systematic review were all designed for use within the context in which they were implemented. Only one study reported a modification where antibiotic prescribing feedback was initially based on the dispensing claim’s date, but in the second year of the study this was modified to reflect the actual prescription date.^26^ Four of the 11 interventions included had a true interactive component where the user could directly manipulate their data and had unlimited access to the dashboard within a defined period of time.^18,19,21,23^ The remaining seven interventions all provided prescribers with longitudinal access to relatively real-time data. Six of the eleven studies included multi-faceted interventions, where the interactive dashboard was part of a broader programme.^18,19,23–25,28^ Two of these were three arm cRCTs where one of the arms received an additional behavioural change component.^23,25^ Two dashboards also alerted prescribers to individual patients with high-risk prescribing/inadequate blood-test monitoring^19^ or inappropriate bronchodilator prescriptions.^27^

**Table 1.**
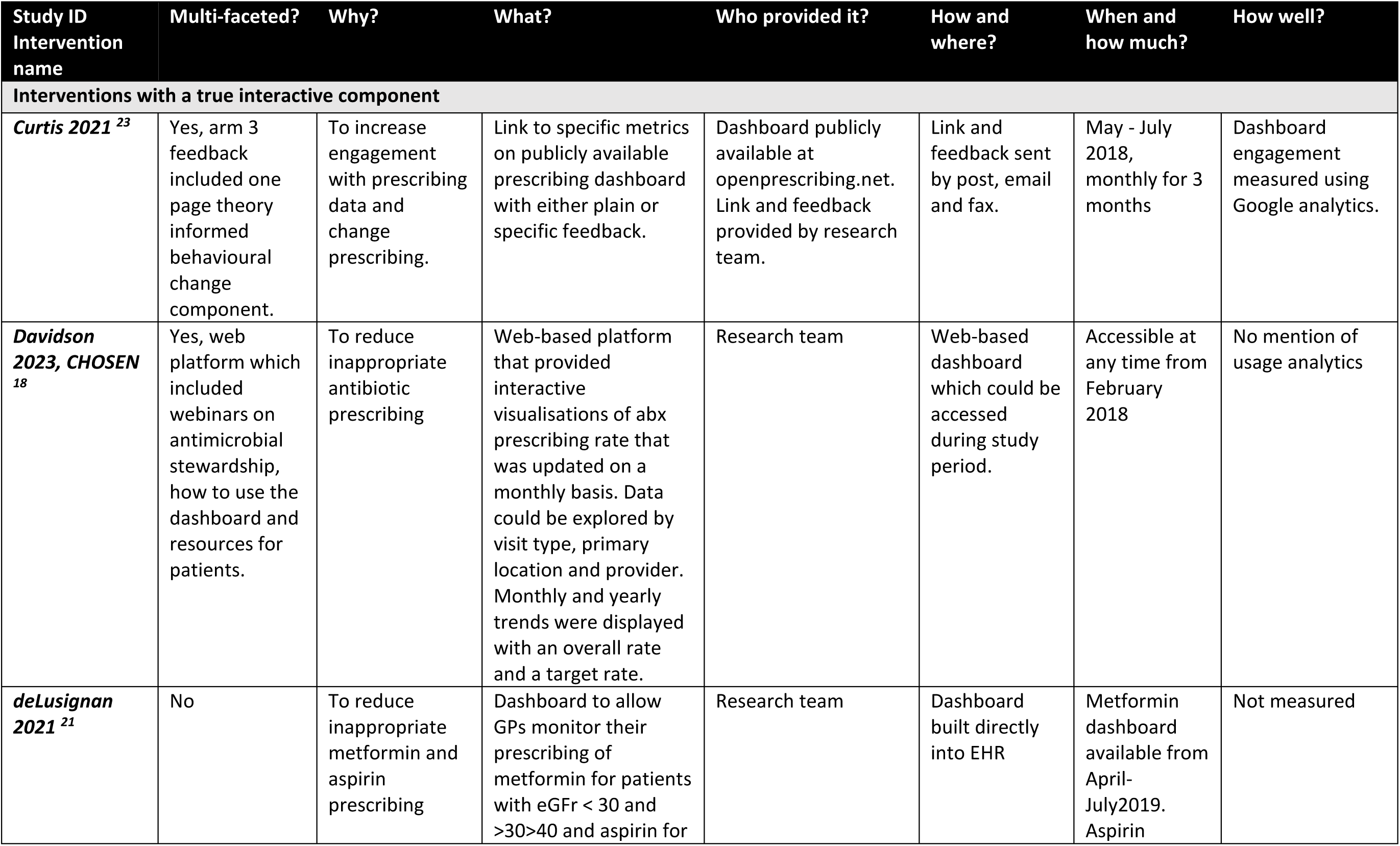
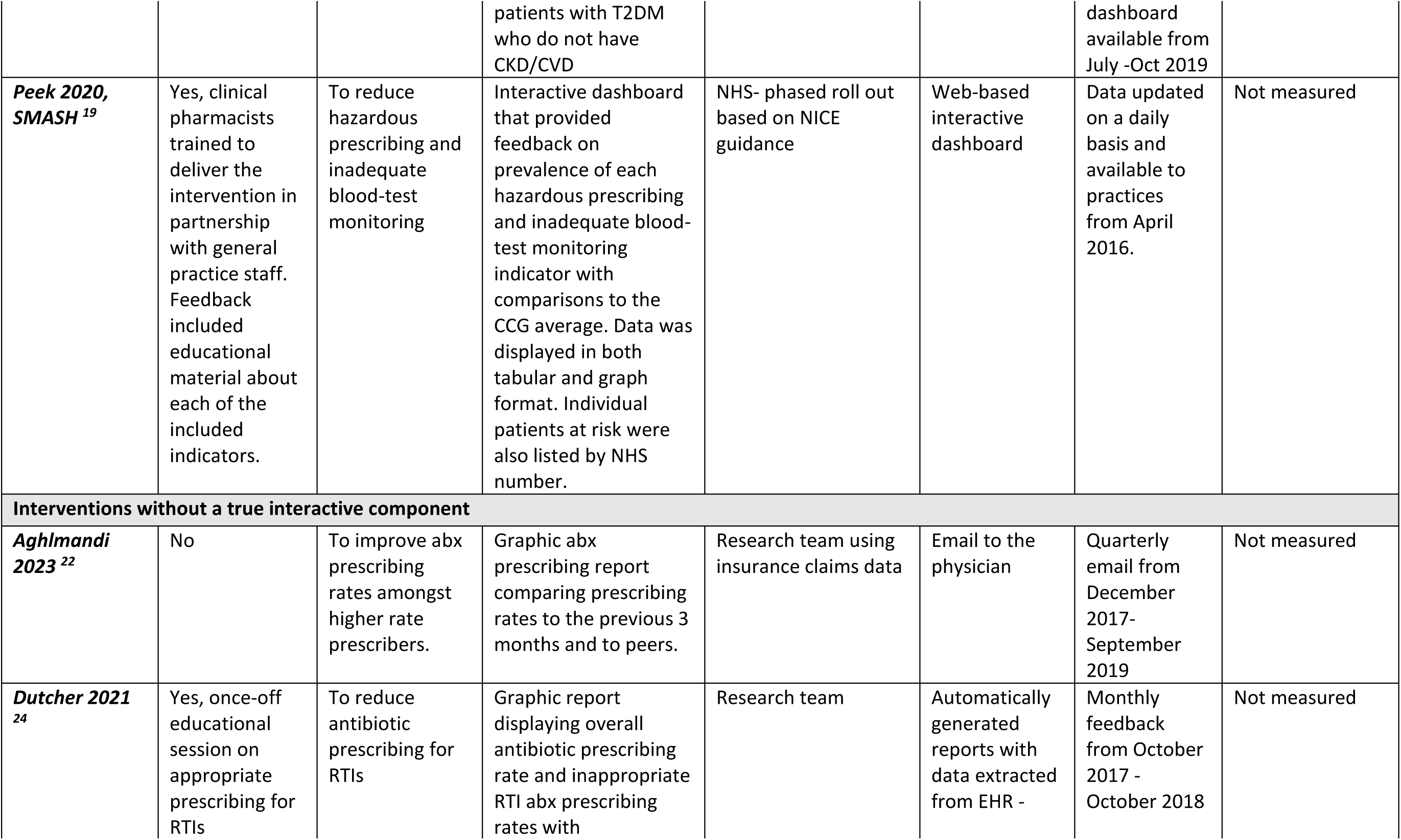
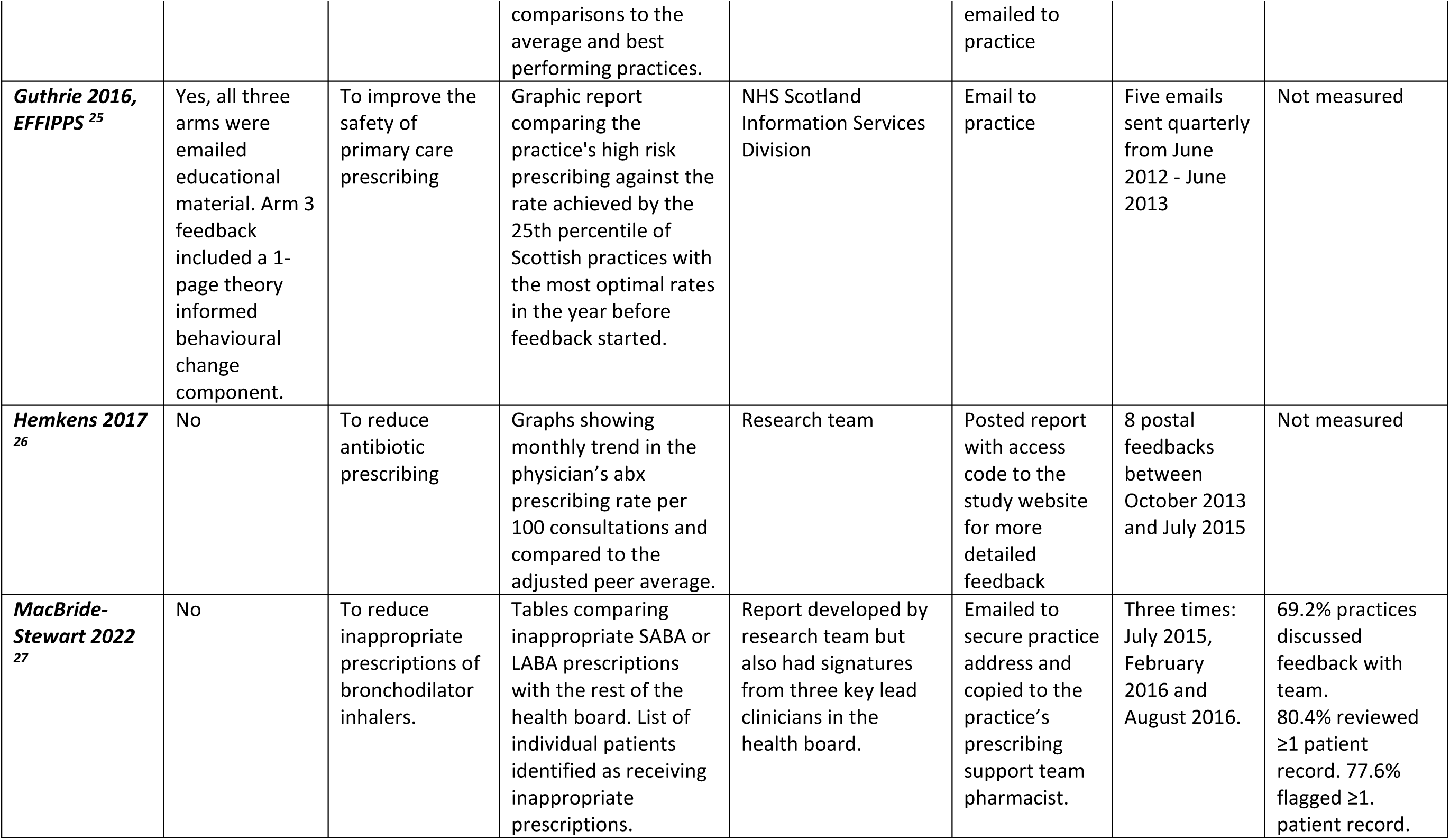
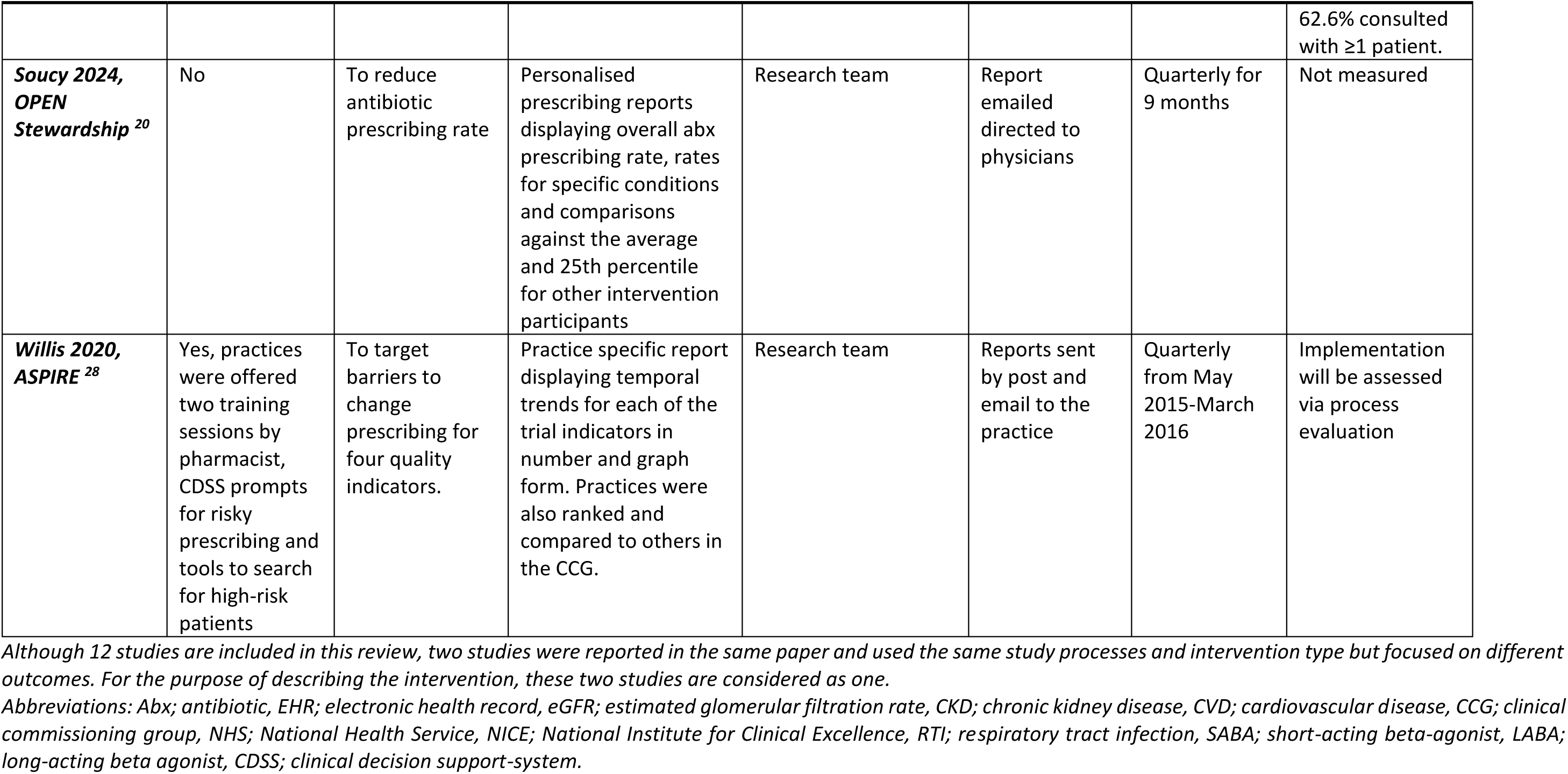
Characteristics of interventions.

### Risk of bias in included studies

Overall nine of the 11 included papers had a low risk of bias,^18,19,22–28^ Figure 2. See S5 Appendix for risk of bias graphs summarising the risk for each EPOC criterion. One ITS had a moderate risk of bias,^20^ where there were significant losses to follow up amongst Canadian physicians that was inadequately addressed, in addition the COVID-19 pandemic was likely to have influenced antibiotic prescribing during the intervention period. There was also insufficient information on prevention of knowledge of allocated interventions and crude post-intervention prescribing rates were not presented which limited transparency.^20^ The included controlled before and after study had a high risk of bias, by virtue of its design.^21^ In addition there was a significant improvement in inappropriate aspirin prescribing prior to intervention implementation, this was clearly reported by the authors’ but unexplained,^21^ given this finding the intervention effect was not considered significant for the purpose of this review.

**Figure 2.**
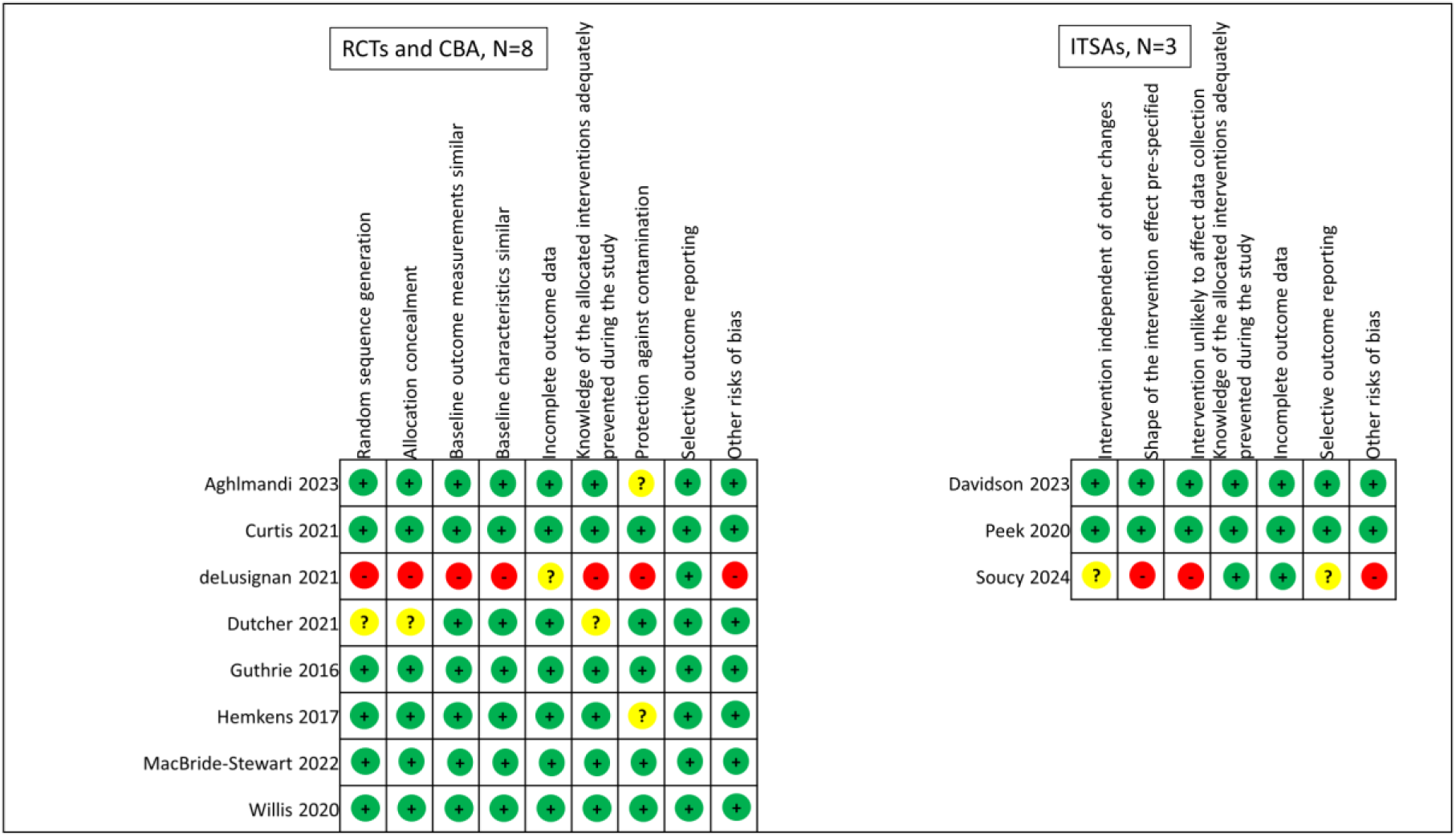
Cochrane Effective Practice and Organisation of Care Risk of bias summary. The two studies reported within the one paper were considered as one for the risk of bias assessment as they had the exact same study processes.

### Effectiveness of interventions

Overall, six of the 12 included studies, two ITSs^18,19^ and four RCTs^24,25,27,28^ demonstrated a significant effect on prescribing-related outcomes, Table 2. When exploring the intervention effect by outcome measure, two of the six studies that targeted antibiotic prescribing had a significant effect.^18,24^ One of these had a true interactive component where prescribers had ongoing access to real-time data^18^ and both were multi-faceted interventions that included an educational component.^18,24^ One was a prospective ITS where practices after the pre-intervention and wash-out periods received access to a web platform that provided educational material and interactive dashboards.^18^ Data was updated on a monthly basis and displayed as graphs illustrating monthly and yearly trends in the antibiotic prescribing rate that could be explored by visit type, primary location and provider.^18^ The effect of the intervention was explored by provider type (paediatric, internal medicine, family physicians and urgent care) with the family physician rate reported here, Table 2. The largest effect size was seen for this group, although significant results were seen in all groups.^18^ The other study that showed a significant effect on antibiotic prescribing was an RCT that focused on inappropriate antibiotic prescribing for respiratory tract infections,^24^ with the outcome measured at the visit level (i.e. the proportion of respiratory tract infection visits that resulted in the prescription of an antibiotic). Participating practices received an educational package and monthly reports by email which displayed the overall antibiotic prescribing rate and the rates of inappropriate antibiotic prescribing for defined respiratory tract infections, both of which were compared to the average and best performing practices.^24^ The remaining four studies that targeted antibiotic prescribing failed to show an effect on antibiotic prescribing rates^20,22,26^ or broad-spectrum antibiotic prescribing.^23^

**Table 2.**
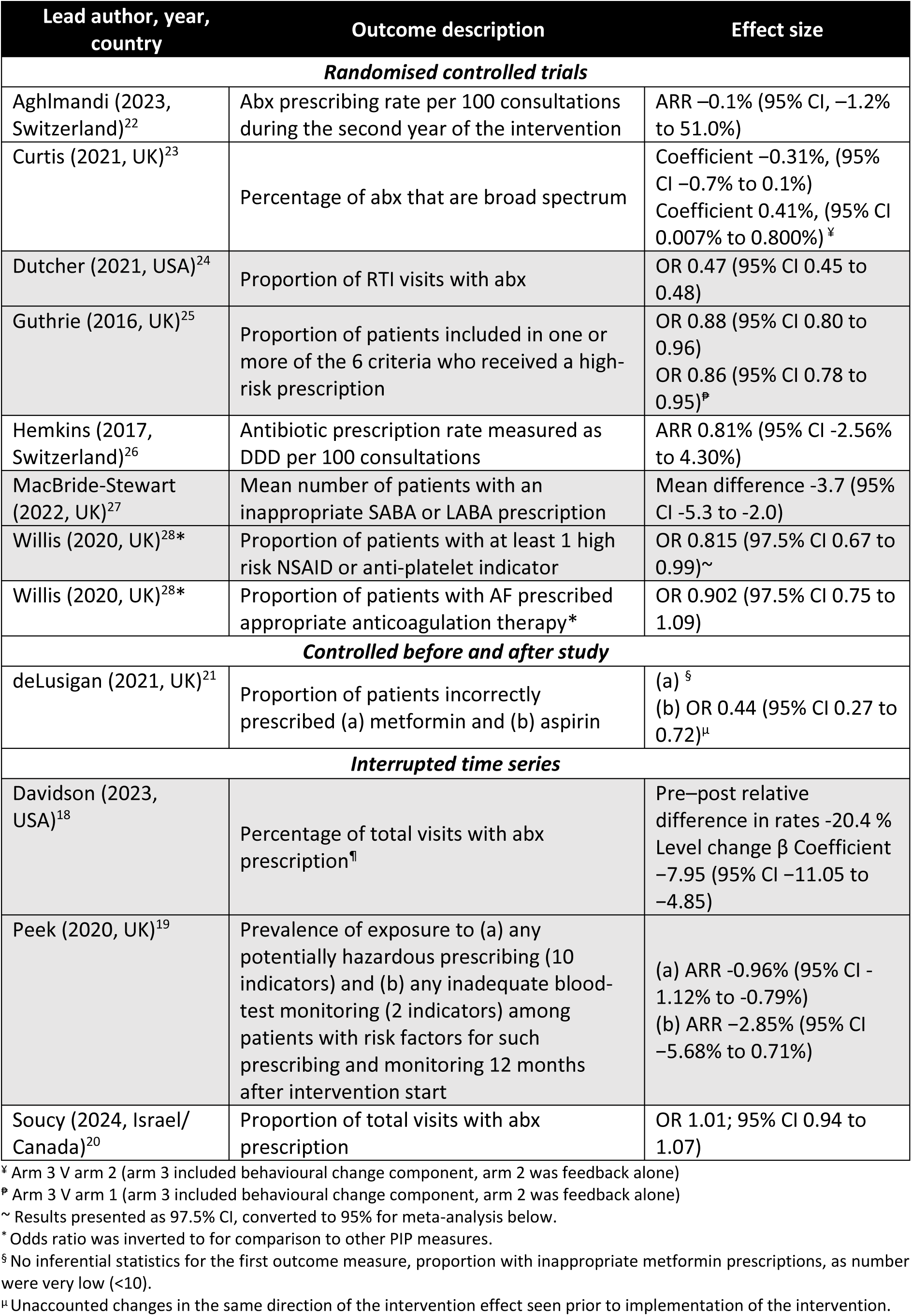
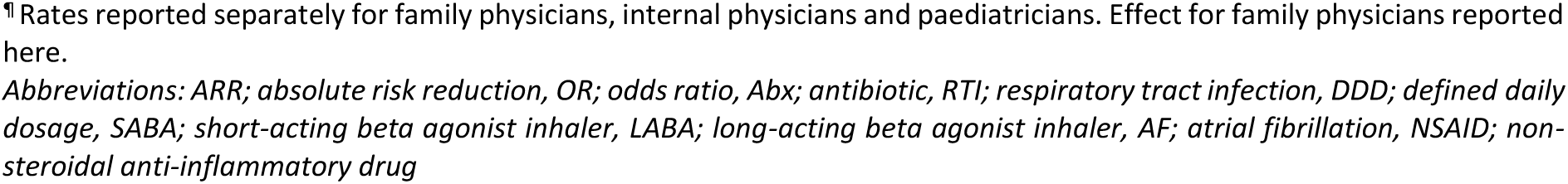
Results of included studies.

Six studies (reported in five papers) explored the effect on PIP for other drugs groups (see S6 Appendix for a list of the indicators included in these studies) and four of these demonstrated a significant effect.^19,25,27,28^ Three of these four were part of multi-faceted interventions that included either a clinical decision support or educational element,^19,25,28^ and only one was a true interactive dashboard where prescribers had ongoing access to real-time data.^19^ The four studies that demonstrated a significant effect on PIP included one ITS and three RCTs. The ITS explored the effect of the routine roll-out of the SMASH intervention that provided feedback on the prevalence of each hazardous prescribing and inadequate blood-test monitoring indicator with comparisons to the local area (clinical commissioning group) average.^19^ There was a significant effect on high-risk prescribing (ARR -0.96%, 95% CI -1.12% to -0.79%) but not on inadequate blood-test monitoring,^19^ however there was a significant effect seen on the latter outcome at 24 weeks follow-up. In addition the largest reductions in high-risk prescribing were seen in practices with higher baseline rates and by 12 months follow-up there was significantly reduced inter-practice variation in the rates of PIP and inadequate blood test monitoring.^19^ The remaining three studies demonstrating a significant effect on PIP were RCTs where feedback with comparative rates of each of the included indicators were emailed to the practice every quarter.^25,27,28^

Of the two studies that did not demonstrate an effect on PIP, one was a controlled before and after study that had a high risk of bias.^21^ Although the authors reported a significant effect on inappropriate aspirin prescribing there was an unexplained significant improvement in this indicator during phase one of the study prior to the implementation of the aspirin dashboard.^21^ In addition no rates or inferential statistics for the effect of the dashboard on inappropriate metformin prescribing were reported as there were only 8 patients prescribed this medicine at follow up.^21^ The other study that did not demonstrate an effect on PIP was a cRCT that explored the effect on inappropriate anticoagulant omissions for atrial fibrillation.^28^

With respect to the intervention components five of the six studies that had multi-faceted interventions showed a significant effect,^18,19,24,25,28^ compared to one^27^ of the five studies that just had the interactive dashboard component. Two cRCTs included clinical decision support elements where individual patients were identified within the dashboard (hazardous prescribing/inadequate blood test monitoring or inappropriate bronchodilator prescribing) and both had a significant effect.^27,28^

Four of the eight included RCTs measured the effect of the intervention on PIP and presented results as proportions and odds ratios with 95% confidence intervals (two of these trials were reported within the one paper).^24,25,28^ However, one of these studies measured the proportion of inappropriate prescriptions at the visit level,^24^ the others were at the patient level.^25,28^ Thus the three RCTs that assessed the effect on appropriate prescribing at the patient level were included in the meta-analysis. The overall effect size was OR 0.91 (95% CI 0.77 to 1.06), with significant heterogeneity (I^2^ =71.8%, p=0.03), see Figure 3. This heterogeneity may be explained by differences in outcome measures: the trial on appropriate atrial fibrillation prescribing assessed improvement in prescribing, whereas the other two studies measured the presence of at least one high-risk prescribing indicator.

**Figure 3.**
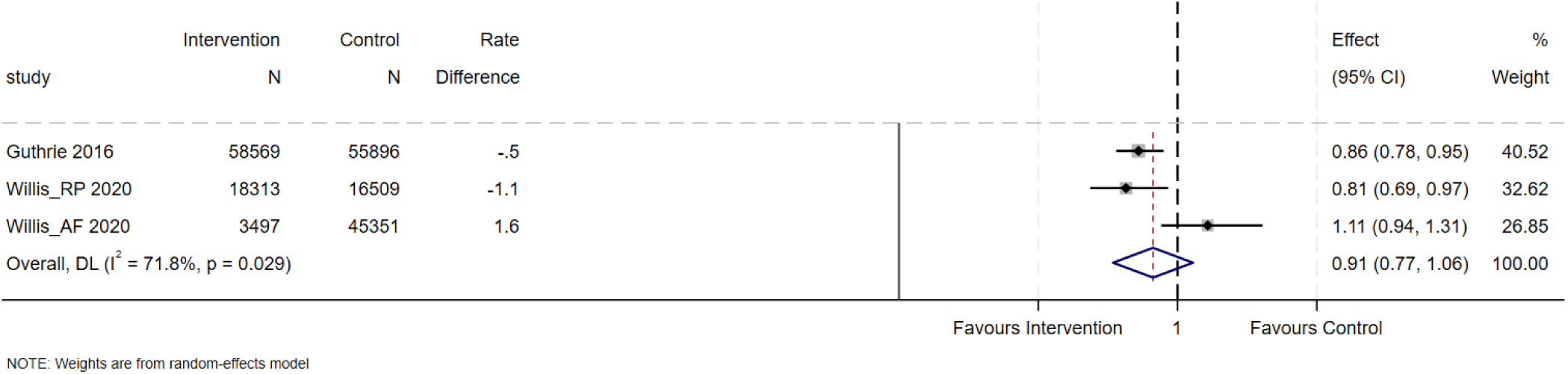
Forest plot of randomised controlled trials exploring the intervention effect on patient level prescribing appropriateness. Abbreviations: CI; confidence interval, RP; risky prescribing, AF; atrial fibrillation Forest plot showing the effects of intervention on prescribing outcomes across included studies. For the two Willis studies, the originally reported 97.5% confidence intervals (CIs) were converted to 95% CIs for consistency. Additionally, for the atrial fibrillation study, the outcome measured was the improvement in appropriate anticoagulant prescribing for atrial fibrillation; the odds ratio (OR) was inverted to align with the direction of effect in the meta-analysis.

## Discussion

### Summary of results

This systematic review aimed to explore the characteristics of interactive dashboard interventions designed to support safe prescribing and explore their effect on prescribing-related outcome measures. Given the nature of the intervention, where data is often fed back on a widespread basis and implemented as a policy, quasi-experimental designs, were included. Six of the 12 included studies demonstrated a significant effect and all of these had an overall low risk of bias.^18,19,24,25,27,28^ Notably, only two of the six studies that explored the effect of these interventions on antibiotic prescribing showed a significant effect.^24^ It may be that recent campaigns to improve antimicrobial stewardship have already resulted in improvements, leaving less room for further advancements. Four of the five studies looking at PIP showed a significant effect.^19,25,27,28^ Potentially inappropriate or high-risk prescribing is more common in complex patients with multimorbidity and polypharmacy^2^ and lack of prescriber awareness has been identified as a barrier to addressing this.^29^ Thus, the difference in effect seen by outcome measure may be because PIP is often related to clinician oversight, whereas antibiotic prescribing reflects a more direct clinician decision and thus may be less amenable to change. This hypothesis is supported by the fact that both interventions that alerted prescribers to specific instances of PIP had a significant effect.^19,27^ Similar to results presented in other systematic reviews exploring the effectiveness of interventions in addressing inappropriate polypharmacy,^30^ multi-faceted interventions were more often effective with five of the six multi-faceted interventions demonstrating a significant effect.^18,19,24,25,28^ In summary, interventions targeting PIP, that included multifaceted elements seemed to have a beneficial effect on outcomes.

Audit and feedback is known to lead to small improvements in professional behaviour, as evidenced by a systematic review of 140 studies, including a meta-analysis of 108 comparisons from 70 studies, which found a median absolute increase of 4.3% in healthcare professionals’ compliance with desired practices.^9^ Increased frequency of feedback, coupled with explicit, measurable targets and specific action plans, were associated with greater effectiveness.^9^ The interventions included in this review all included ongoing feedback of relatively contemporaneous data, with comparisons to specific set targets or comparative benchmarking. There was significant heterogeneity among the results of the three studies included in this meta-analysis. This was likely explained by the differing outcome measure in the study assessing the effect on appropriate coagulation for atrial fibrillation, where the outcome reflected inappropriate omission as opposed to an inappropriate prescription. Other approaches such educational interventions and clinical decision support have also failed to improve anticoagulation rates for patients with atrial fibrillation.^31^ Collaborative, shared decision making interventions may be needed to address this more complex prescribing decision around reducing the future risk of an adverse event.^32^ The two other studies included in the meta-analysis, both of which demonstrated a significant effect, targeted PIP and included an educational component,^25,28^ which may have mediated some of the observed effect. The study by Willis et al, which showed a larger treatment effect, also incorporated clinical decision support, which has been shown to have a modest effect on prescribing behaviour.^33^

### Strengths and limitations

This systematic review included a broad and detailed search strategy including citation chasing and grey literature searches of clinical trial registries to reduce the risk of publication bias. Including quasi-experimental designs meant we captured two large ITSs that explored the implementation of novel interactive dashboards. Given the heterogeneous nature of the outcome measures included (different prescribing criteria, measured at different levels (e.g. practice and visit level as well as patient level) it was only possible to perform a meta-analysis for three studies and it was not possible to conduct a funnel plot to formally assess publication bias. The scope of our review was shaped by focusing on feedback mechanisms that align with the functionality of interactive dashboards. While this approach excludes one-off feedback interventions, it allows for a more detailed exploration of tools designed for repeated engagement with contemporaneous data, which are hypothesised to have distinct advantages.^34^ This focus also aligns with prior evidence suggesting that repeated feedback may be more effective in driving behaviour change,^35^ though we acknowledge that findings from recent research indicate variability in this relationship.^36^

### Implications for research, policy and practice

This review identified several gaps in the existing literature. Only 12 studies met the inclusion criteria, with just four employing true interactive dashboards and six were multifaceted. The limited number of studies suggests a need for further research exploring the effectiveness of interactive dashboards designed to optimise prescribing. A key challenge in conducting the meta-analysis was the heterogeneity of outcome measures. Identifying appropriate outcome measures is a challenge when assessing the effectiveness of interventions such as interactive dashboards designed to improve prescribing quality in primary care settings. Important clinical endpoints such as unplanned hospital admissions or mortality require a sufficient sample size and an adequate follow-up period to identify any potential effect mediated by higher quality prescribing, which may not be feasible. Thus, composite measures of explicit prescribing criteria are often used as primary endpoints for these studies.^37^ There is strong observational evidence that such prescribing is associated with adverse outcomes for patients such as increased mortality, falls and unplanned hospital admissions.^38^ More general prescribing measures include rates of utilisation and these serve as a valuable outcome for evaluating the impact of health policy implementation programmes aimed at modifying prescribing behaviours, such as reducing high opioid prescription levels^39^ or imposing restrictions on drugs with unfavourable cost-effectiveness profiles.^40^ However, the outcomes identified for this review were reported at various levels, including the prescription, patient, prescriber, and practice levels. The development of a core outcome set for prescribing-related measures that can be consistently applied across studies utilising routine prescribing or dispensing datasets may be one way of addressing this and would facilitate more meaningful comparisons and benefit future meta-analyses. Future research should also investigate how different feedback features such as frequency, content, and format influence both effectiveness and engagement, and whether there is an optimal level of engagement required for behaviour change. This will necessitate systems capable of providing both ongoing feedback and measuring user engagement through detailed use analytics, the latter of which was notably absent in most studies included in this systematic review. Finally, routine evaluations of policies that utilise this data to optimise prescribing are essential. Systematic and standardised evaluations would provide valuable insights into the effectiveness of policy interventions aimed at optimising prescribing, thereby enhancing clinical outcomes and health system efficiency.

## Conclusion

Interactive dashboards have the potential to support safe and effective prescribing in primary care. Multi-faceted interventions that target high-risk prescribing are more likely to be effective. Future research should focus on developing core outcome sets to facilitate future meta-analyses of effectiveness as well as optimising their implementation and understanding how to sustain user engagement. To support their implementation, it is essential to establish the necessary data infrastructure within primary care systems. With advancements in data infrastructure and analysis, these interventions could have a significant impact if implemented at scale.

## Data Availability

All data produced in the present work are contained in the manuscript

## Acknowledgments

Killian Walsh; Information Specialist, RCSI library assisted with search strategy. Mobeena Naz; Medical Student, RCSI assisted with title and abstract screening.

## Author roles

**Caroline McCarthy:** Conceptualization, Formal Analysis, Funding Acquisition, Investigation, Methodology, Project Administration, Resources, Supervision, Validation, Visualization, Writing – Original Draft Preparation, Writing – Review & Editing

**Patrick Moynagh:** Investigation, Methodology, Project Administration, Resources, Supervision, Writing – Review & Editing

**Aine Mannion:** Investigation, Writing – Review & Editing

**Ashley Wei**: Investigation, Writing – Review & Editing

**Barbara Clyne:** Conceptualization, Funding Acquisition, Methodology, Writing – Review & Editing

**Frank Moriarty:** Conceptualization, Funding Acquisition, Investigation, Methodology, Supervision, Validation, Writing – Review & Editing

## Data Availability

No new primary data were collected for this study, as the review synthesises publicly available data from published studies. All data generated or analysed during this study are included in this published article and its supplementary information files. This includes the search strategies used for each database, details of the included studies, and any additional data extracted during the review process. The study protocol is available as an open-access publication and can be accessed directly at https://hrbopenresearch.org/articles/7-44.

## Abbreviations

PIP: potentially inappropriate prescribing
EHR: electronic healthcare records
RCT: randomised controlled trial
ITS: interrupted time series
CBA: controlled before and after study
EPOC: Effective Practice and Organisation of Care
TIDieR: Template for Intervention Description and Replication
Abx: antibiotic
SABA: short acting beta agonist inhaler
LABA: long acting beta agonist inhaler
NSAID: non-steroidal anti-inflammatory drug
eGFR: estimated glomerular filtration rate
CKD: chronic kidney disease
CVD: cardiovascular disease
CCG: clinical commissioning group
NICE: National Institute for Clinical Excellence
RTI: respiratory tract infection
CDSS: clinical decision support-system
ARR: absolute risk reduction
OR: odds ratio
DDD: defined daily dosage
CI: confidence interval
RP: risky prescribing
AF: atrial fibrillation

## Supplemental material

S1 Appendix: PRISMA Checklist

S2 Appendix: Electronic search reports

S3 Appendix Study Inclusion Criteria

S4 Appendix: List of all data points extracted from included studies

S6 Appendix Characteristics of Included Studies

S7 Appendix: Risk of bias graphs

S8 Appendix: List of all endpoints for included studies

## References

1. Fortin M, Lapointe L, Hudon C, Vanasse A, Ntetu AL, Maltais D. Multimorbidity and quality of life in primary care: a systematic review. Health and Quality of Life Outcomes. 2004;2:51.

2. Pérez T, Moriarty F, Wallace E, McDowell R, Redmond P, Fahey T. Prevalence of potentially inappropriate prescribing in older people in primary care and its association with hospital admission: longitudinal study. BMJ. 2018;363:k4524.

3. Wallace E, Salisbury C, Guthrie B, Lewis C, Fahey T, Smith SM. Managing patients with multimorbidity in primary care. BMJ. 2015;350:h176.

4. Jung Yin Tsang MS, Thomas Blakeman, Rupert Payne, Darren M Aschcroft. Protocol for the development and validation of a Polypharmacy Assessment Score for potentially problematic polypharmacy PREPRINT (Version 1) available at Research Square 2023.

5. Chang CB, Chan DC. Comparison of published explicit criteria for potentially inappropriate medications in older adults. Drugs Aging. 2010;27(12):947–957.

6. Radomski TR, Decker A, Khodyakov D, et al. Development of a Metric to Detect and Decrease Low-Value Prescribing in Older Adults. JAMA network open. 2022;5(2):e2148599–e2148599.

7. Dimitrow MS, Airaksinen MS, Kivela SL, Lyles A, Leikola SN. Comparison of prescribing criteria to evaluate the appropriateness of drug treatment in individuals aged 65 and older: a systematic review. J Am Geriatr Soc. 2011;59(8):1521–1530.

8. Curtis HJ, Goldacre B. OpenPrescribing: normalised data and software tool to research trends in English NHS primary care prescribing 1998–2016. 2018;8(2):e019921.

9. Ivers N, Jamtvedt G, Flottorp S, et al. Audit and feedback: effects on professional practice and healthcare outcomes. Cochrane Database Syst Rev. 2012(6):Cd000259.

10. Kwan JL, Lo L, Ferguson J, et al. Computerised clinical decision support systems and absolute improvements in care: meta-analysis of controlled clinical trials. 2020;370:m3216.

11. Williams R, Keers R, Gude WT, et al. SMASH! The Salford medication safety dashboard. 2018;25(3):183–193.

12. Moynagh P, Mannion, Á, Wei, A, Clyne, B, Moriarty, F, McCarthy, C. Effectiveness of interactive dashboards to optimise prescribing in primary care: a protocol for a systematic review [version 1; peer review: awaiting peer review]. HRB open research. 2024;7(44).

13. Higgins JPT, Thomas J, Chandler J, et al. Cochrane Handbook for Systematic Reviews of Interventions version 6.4 (updated August 2023). In: Cochrane; 2023: www.training.cochrane.org/handbook.

14. Page MJ, McKenzie JE, Bossuyt PM, et al. The PRISMA 2020 statement: an updated guideline for reporting systematic reviews. BMJ. 2021:n71.

15. Haddaway NR, Grainger MJ, Gray CT. Citationchaser: A tool for transparent and efficient forward and backward citation chasing in systematic searching. Research synthesis methods. 2022;13(4):533–545.

16. Cochrane Effective Practice and Organisation of Care (EPOC). What study designs can be considered for inclusion in an EPOC review and what should they be called?. 2017. https://epoc.cochrane.org/sites/epoc.cochrane.org/files/uploads/EPOC%20Study%20Designs%20About.pdf. Accessed 13/03/2024.

17. Hoffmann TC, Glasziou PP, Boutron I, et al. Better reporting of interventions: template for intervention description and replication (TIDieR) checklist and guide. 2014;348:g1687.

18. Davidson LE, Gentry EM, Priem JS, Kowalkowski M, Spencer MD. A multimodal intervention to decrease inappropriate outpatient antibiotic prescribing for upper respiratory tract infections in a large integrated healthcare system. Infect Control Hosp Epidemiol. 2023;44(3):392–399.

19. Peek N, Gude WT, Keers RN, et al. Evaluation of a pharmacist-led actionable audit and feedback intervention for improving medication safety in UK primary care: An interrupted time series analysis. PLoS Med. 2020;17(10):1–17.

20. Soucy Jean-Paul R, Low M, Acharya Kamal R, et al. Evaluation of an automated feedback intervention to improve antibiotic prescribing among primary care physicians (OPEN Stewardship): a multinational controlled interrupted time-series study. Microbiology spectrum. 2024;12(4):e00017–00024.

21. de Lusignan S, Hinton W, Seidu S, et al. Dashboards to reduce inappropriate prescribing of metformin and aspirin: A quality assurance programme in a primary care sentinel network. Prim Care Diabetes. 2021;15(6):1075–1079.

22. Aghlmandi S, Halbeisen FS, Saccilotto R, et al. Effect of Antibiotic Prescription Audit and Feedback on Antibiotic Prescribing in Primary Care: A Randomized Clinical Trial. JAMA internal medicine. 2023;183(3):213–220.

23. Curtis HJ, Bacon S, Croker R, et al. Evaluating the impact of a very low-cost intervention to increase practices’ engagement with data and change prescribing behaviour: a randomized trial in English primary care. Fam Pract. 2021;38(4):373–380.

24. Dutcher L, Degnan KO, Adu-Gyamfi AB, et al. Improving Outpatient Antibiotic Prescribing for Respiratory Tract Infections in Primary Care; a Stepped-Wedge Cluster Randomized Trial. Clinical infectious diseases: an official publication of the Infectious Diseases Society of America. 2021;74(6):947–956.

25. Guthrie B, Kavanagh K, Robertson C, et al. Data feedback and behavioural change intervention to improve primary care prescribing safety (EFIPPS): multicentre, three arm, cluster randomised controlled trial. BMJ (Clinical research ed). 2016;354:i4079.

26. Hemkens LG, Saccilotto R, Leon Reyes S, et al. Personalized Prescription Feedback Using Routinely Collected Data to Reduce Antibiotic Use in Primary Care: A Randomized Clinical Trial. JAMA internal medicine. 2017;177(2):176–183.

27. MacBride-Stewart S, Marwick C, Ryan M, Guthrie B. Feedback of actionable individual patient prescription data to improve asthma prescribing: pragmatic cluster randomised trial in 233 UK general practices. Br J Gen Pract. 2022;72(722):e627–633.

28. Willis TA, Collinson M, Glidewell L, et al. An adaptable implementation package targeting evidence-based indicators in primary care: A pragmatic cluster-randomised evaluation. PLoS Med. 2020;17(2):1–20.

29. Anderson K, Stowasser D, Freeman C, Scott I. Prescriber barriers and enablers to minimising potentially inappropriate medications in adults: a systematic review and thematic synthesis. BMJ open. 2014;4(12).

30. Clyne B, Fitzgerald C, Quinlan A, et al. Interventions to Address Potentially Inappropriate Prescribing in Community-Dwelling Older Adults: A Systematic Review of Randomized Controlled Trials. J Am Geriatr Soc. 2016;64(6):1210–1222.

31. Kapoor A, Amroze A, Vakil F, et al. SUPPORT-AF II: Supporting Use of Anticoagulants Through Provider Profiling of Oral Anticoagulant Therapy for Atrial Fibrillation: A Cluster-Randomized Study of Electronic Profiling and Messaging Combined With Academic Detailing for Providers Making Decisions About Anticoagulation in Patients With Atrial Fibrillation. Circ Cardiovasc Qual Outcomes. 2020;13(2):e005871.

32. Siontis KC, Montori VM, Noseworthy PA. Multimodal Interventions to Increase Anticoagulant Utilization in Atrial Fibrillation: Futile Without Patient Engagement? Circ Cardiovasc Qual Outcomes. 2020;13(2):e006418.

33. Shojania KG, Jennings A, Mayhew A, Ramsay CR, Eccles MP, Grimshaw J. The effects of on-screen, point of care computer reminders on processes and outcomes of care. Cochrane Database Syst Rev. 2009;2009(3):Cd001096.

34. Colquhoun HL, Carroll K, Eva KW, et al. Advancing the literature on designing audit and feedback interventions: identifying theory-informed hypotheses. Implementation Science. 2017;12(1):117.

35. Ivers NM, Grimshaw JM, Jamtvedt G, et al. Growing literature, stagnant science? Systematic review, meta-regression and cumulative analysis of audit and feedback interventions in health care. J Gen Intern Med. 2014;29(11):1534–1541.

36. Schwartz KL, Shuldiner J, Langford BJ, et al. Mailed feedback to primary care physicians on antibiotic prescribing for patients aged 65 years and older: pragmatic, factorial randomised controlled trial. BMJ. 2024;385:e079329.

37. Cole JA, Gonçalves-Bradley DC, Alqahtani M, et al. Interventions to improve the appropriate use of polypharmacy for older people. Cochrane Database Syst Rev. 2023(10).

38. Mekonnen AB, Redley B, de Courten B, Manias E. Potentially inappropriate prescribing and its associations with health-related and system-related outcomes in hospitalised older adults: A systematic review and meta-analysis. Br J Clin Pharmacol. 2021;87(11):4150–4172.

39. Daoust R, Paquet J, Marquis M, et al. Evaluation of Interventions to Reduce Opioid Prescribing for Patients Discharged From the Emergency Department: A Systematic Review and Meta-analysis. JAMA network open. 2022;5(1):e2143425–e2143425.

40. Mattsson M, Boland F, Kirke C, et al. The impact of lidocaine plaster prescribing reduction strategies: A comparison of two national health services in Europe. Br J Clin Pharmacol. 2023;89(8):2349–2358.

