## Supplemental material for "Effectiveness of interactive dashboards to optimise prescribing in general practice: A systematic review"

#### S1 Appendix: PRISMA Checklist

| Section and Topic | Item # | Checklist item | Location where item is reported |
|---|---|---|---|
| **TITLE** | | | |
| Title | 1 | Identify the report as a systematic review. | Title page |
| **ABSTRACT** | | | |
| Abstract | 2 | See the PRISMA 2020 for Abstracts checklist. | Abstract |
| **INTRODUCTION** | | | |
| Rationale | 3 | Describe the rationale for the review in the context of existing knowledge. | Introduction, page 3 |
| Objectives | 4 | Provide an explicit statement of the objective(s) or question(s) the review addresses. | Introduction, page 4 |
| **METHODS** | | | |
| Eligibility criteria | 5 | Specify the inclusion and exclusion criteria for the review and how studies were grouped for the syntheses. | Methods, page 5-6 |
| Information sources | 6 | Specify all databases, registers, websites, organisations, reference lists and other sources searched or consulted to identify studies. Specify the date when each source was last searched or consulted. | Methods, page 5 |
| Search strategy | 7 | Present the full search strategies for all databases, registers and websites, including any filters and limits used. | Appendix 2 |
| Selection process | 8 | Specify the methods used to decide whether a study met the inclusion criteria of the review, including how many reviewers screened each record and each report retrieved, whether they worked independently, and if applicable, details of automation tools used in the process. | Methods, page 6 |
| Data collection process | 9 | Specify the methods used to collect data from reports, including how many reviewers collected data from each report, whether they worked independently, any processes for obtaining or confirming data from study investigators, and if applicable, details of automation tools used in the process. | Methods, page 6 |
| Data items | 10a | List and define all outcomes for which data were sought. Specify whether all results that were compatible with each outcome domain in each study were sought (e.g. for all measures, time points, analyses), and if not, the methods used to decide which results to collect. | Methods, page 6, Appendix 3 |
| | 10b | List and define all other variables for which data were sought (e.g. participant and intervention characteristics, funding sources). Describe any assumptions made about any missing or unclear information. | Methods, analysis, page 6 |
| Study risk of bias assessment | 11 | Specify the methods used to assess risk of bias in the included studies, including details of the tool(s) used, how many reviewers assessed each study and whether they worked independently, and if applicable, details of automation tools used in the process. | Methods, page 6 |
| Effect measures | 12 | Specify for each outcome the effect measure(s) (e.g. risk ratio, mean difference) used in the synthesis or presentation of results. | Results, Table 3 |
| Synthesis methods | 13a | Describe the processes used to decide which studies were eligible for each synthesis (e.g. tabulating the study intervention characteristics and comparing against the planned groups for each synthesis (item #5)). | Methods, analysis, page 6 |
| | 13b | Describe any methods required to prepare the data for presentation or synthesis, such as handling of missing summary statistics, or data conversions. | Methods, analysis, page 6 |
| | 13c | Describe any methods used to tabulate or visually display results of individual studies and syntheses. | Methods, analysis, page 6 |
| | 13d | Describe any methods used to synthesize results and provide a rationale for the choice(s). If meta-analysis was performed, describe the model(s), method(s) to identify the presence and extent of statistical heterogeneity, and software package(s) used. | Methods, analysis, page 6 |
| | 13e | Describe any methods used to explore possible causes of heterogeneity among study results (e.g. subgroup analysis, meta-regression). | N/A |
| | 13f | Describe any sensitivity analyses conducted to assess robustness of the synthesized results. | N/A |
| Reporting bias assessment | 14 | Describe any methods used to assess risk of bias due to missing results in a synthesis (arising from reporting biases). | Page 20 |

| Section and Topic | Item # | Checklist item | Location where item is reported |
|---|---|---|---|
| Certainty assessment | 15 | Describe any methods used to assess certainty (or confidence) in the body of evidence for an outcome. | Methods, analysis, page 6 |
| **RESULTS** | | | |
| Study selection | 16a | Describe the results of the search and selection process, from the number of records identified in the search to the number of studies included in the review, ideally using a flow diagram. | Figure 1 |
| | 16b | Cite studies that might appear to meet the inclusion criteria, but which were excluded, and explain why they were excluded. | Appendix 4 |
| Study characteristics | 17 | Cite each included study and present its characteristics. | Results, Table 1 |
| Risk of bias in studies | 18 | Present assessments of risk of bias for each included study. | Results, Figure 2 |
| Results of individual studies | 19 | For all outcomes, present, for each study: (a) summary statistics for each group (where appropriate) and (b) an effect estimate and its precision (e.g. confidence/credible interval), ideally using structured tables or plots. | Results, Table 3 |
| Results of syntheses | 20a | For each synthesis, briefly summarise the characteristics and risk of bias among contributing studies. | Results,<br>Page 15,16 |
| | 20b | Present results of all statistical syntheses conducted. If meta-analysis was done, present for each the summary estimate and its precision (e.g. confidence/credible interval) and measures of statistical heterogeneity. If comparing groups, describe the direction of the effect. | Results, page 18 |
| | 20c | Present results of all investigations of possible causes of heterogeneity among study results. | Results, Page 18 |
| | 20d | Present results of all sensitivity analyses conducted to assess the robustness of the synthesized results. | N/A |
| Reporting biases | 21 | Present assessments of risk of bias due to missing results (arising from reporting biases) for each synthesis assessed. | Discussion, Page 20 |
| Certainty of evidence | 22 | Present assessments of certainty (or confidence) in the body of evidence for each outcome assessed. | Table 3, Figure 3 |
| **DISCUSSION** | | | |
| Discussion | 23a | Provide a general interpretation of the results in the context of other evidence. | Page 19 |
| | 23b | Discuss any limitations of the evidence included in the review. | Page 20 |
| | 23c | Discuss any limitations of the review processes used. | Page 20 |
| | 23d | Discuss implications of the results for practice, policy, and future research. | Page 20-21 |
| **OTHER INFORMATION** | | | |
| Registration and protocol | 24a | Provide registration information for the review, including register name and registration number, or state that the review was not registered. | Methods, Page 5 |
| | 24b | Indicate where the review protocol can be accessed, or state that a protocol was not prepared. | Methods, Page 5 |
| | 24c | Describe and explain any amendments to information provided at registration or in the protocol. | No amendments |
| Support | 25 | Describe sources of financial or non-financial support for the review, and the role of the funders or sponsors in the review. | Funding statement |
| Competing interests | 26 | Declare any competing interests of review authors. | COI statement |
| Availability of data, code and other materials | 27 | Report which of the following are publicly available and where they can be found: template data collection forms; data extracted from included studies; data used for all analyses; analytic code; any other materials used in the review. | Supplementary material |

*From:* Page MJ, McKenzie JE, Bossuyt PM, Boutron I, Hoffmann TC, Mulrow CD, et al. The PRISMA 2020 statement: an updated guideline for reporting systematic reviews. BMJ 2021;372:n71. doi: 10.1136/bmj.n71

#### S2 Appendix: Electronic search reports

| Electronic search report No. 1 | |
|---|---|
| Electronic database | Ovid MEDLINE(R) and Epub Ahead of Print, In-Process, In-Data-Review & Other Non-Indexed Citations, Daily and Versions 1946 to 17th August, 2023 |
| Platform | OVID |
| Date of search | 17th November 2023 |
| Range of date | None |
| Restriction of language | None |
| Other limits | None |
| Search Strategy | 1. exp general Practitioners/<br>2. general practice physician*.tw.<br>3. general practitioner*.tw.<br>4. GPs.tw.<br>5. exp Family Practice/ or general practice.tw.<br>6. (family and (practice* or doctor*)).tw.<br>7. (Clinician* or physician*).tw.<br>8. primary care practice*.tw.<br>9. primary care.tw.<br>10. exp Primary Health Care/<br>11. 1 or 2 or 3 or 4 or 5 or 6 or 7 or 8 or 9 or 10<br>12. exp electronic Health Records/<br>13. ((electronic or computeri#ed) and (health or medical) and record*).tw.<br>14. exp User-Computer Interface/ or computer interface.tw.<br>15. (('user computer' or user) adj3 computer).tw.<br>16. Dashboard*.tw.<br>17. visual analytic*.tw.<br>18. exp Health Information System/ or medical information system.tw.<br>19. Health Information System* Data.tw.<br>20. Virtual Desktop Infrastructure.tw.<br>21. data visualisation*.tw.<br>22. exp Computer Graphics/<br>23. computer graphic*.tw.<br>24. exp clinical audit/ and exp medical audit/<br>25. benchmarking.tw.<br>26. feedback.tw.<br>27. visual health record.tw.<br>28. Interactive visualization tool*.tw.<br>29. ((paperless or digital or electronic or online) and (health data or healthcare system* or health care system or information processing or personalised prescription* or personalized prescription*)).tw.<br>30. 12 or 13 or 14 or 15 or 16 or 17 or 18 or 19 or 20 or 21 or 22 or 23 or 24 or 25 or 26 or 27 or 28 or 29<br>31. 11 and 30<br>32. exp drug Prescriptions/<br>33. (Drug and Prescri*).tw.<br>34. exp inappropriate Prescribing/ or prescribing error.tw.<br>35. ((inappropriate or over) adj3 prescri*).tw.<br>36. (reduc* and prescription*).tw. |

| | |
|---|---|
| | 37. 32 or 33 or 34 or 35 or 36<br>38. 31 and 37 |
| Number of references found | 2595 |

| Electronic search report No. 2 | |
|---|---|
| Electronic database | EBM Reviews – Cochrane Central register of Controlled Trials |
| Platform | OVID |
| Date of search | 17th November 2023 |
| Range of date | None |
| Restriction of language | None |
| Other limits | None |
| Search Strategy | 1. "general practitioner*".tw.<br>2. "general practice physician*".tw.<br>3. GPs.tw.<br>4. ("Family Practice" or "general practice").tw.<br>5. (family and (practice* or doctor*)).tw.<br>6. (Clinician* or physician*).tw.<br>7. "primary care practice*".tw.<br>8. "Primary Health Care".tw.<br>9. "Primary Care".tw.<br>10. 1 or 2 or 3 or 4 or 5 or 6 or 7 or 8 or 9<br>11. "electronic health record*".tw.<br>12. ((electronic or computeri#ed) adj3 (health or medical) adj3 record*).tw.<br>13. ("User-Computer Interface*" or "computer interface*").tw.<br>14. (("User-computer" or "User computer*") adj4 Interface*).tw.<br>15. Dashboard*.tw.<br>16. visual analytic*.tw.<br>17. ("Health Information System*" or "medical information system*").tw.<br>18. Health Information System* Data.tw.<br>19. "Virtual Desktop Infrastructure".tw.<br>20. "data visualisation*".tw.<br>21. "computer graphic*".tw.<br>22. clinical audit.tw.<br>23. "medical audit*".tw.<br>24. "feedback".tw.<br>25. "benchmarking".tw.<br>26. "visual health record*".tw.<br>27. "Interactive visualization tool*".tw.<br>28. (("paperless" or "digital" or "electronic" or "online") and ("health data" or "healthcare system*" or "health care system*" or "information processing" or "personalised prescription*" or "personalized prescription*")).tw.<br>29. 11 or 12 or 13 or 14 or 15 or 16 or 17 or 18 or 19 or 20 or 21 or 22 or 23 or 24 or 25 or 26 or 27 or 28<br>30. 10 and 29<br>31. drug Prescriptions.tw.<br>32. (Drug and Prescri*).tw.<br>33. ("inappropriate Prescribing" or "prescribing error").tw.<br>34. ((Inappropriate or over) and Prescri*).tw.<br>35. (reduc* and prescription*).tw. |

| | |
|---|---|
| | 36. 31 or 32 or 33 or 34 or 35<br>37. 30 and 36 |
| Number of references found | 794 |

| Electronic search report No. 3 | |
|---|---|
| Electronic database | Embase |
| Platform | Elsevier |
| Date of search | 17th November 2023 |
| Range of date | None |
| Restriction of language | None |
| Other limits | |
| Search Strategy | #1 'general practitioner'/exp<br>#2 'general practitioner*':ab,ti<br>#3 'general practice physician*':ab,ti<br>#4 'gps':ab,ti<br>#5 'general practice'/exp<br>#6 'family practice':ab,ti<br>#7 family:ab,ti AND (practice*:ab,ti OR doctor*:ab,ti)<br>#8 clinician*:ti,ab<br>#9 physican*:ti,ab<br>#10 'primary health care'/exp<br>#11 'primary care':ti,ab<br>#12 #1 OR #2 OR #3 OR #4 OR #5 OR #6 OR #7 OR #8 OR #9 OR #10 OR #11<br>#13 'electronic health record'/exp<br>#14 (electronic:ab,ti OR computeri?ed:ab,ti) AND (health:ab,ti OR medical:ab,ti) AND record*:ab,ti<br>#15 'computer interface'/exp<br>#16 'user-computer interface':ab,ti<br>#17 (('user computer' OR user) NEXT/3 computer):ti,ab<br>#18 dashboard*:ab,ti<br>#19 'visual analytics':ab,ti<br>#20 'medical information system'/exp<br>#21 'health information system':ab,ti<br>#22 'health information system* data':ab,ti<br>#23 'virtual desktop infrastructure':ab,ti<br>#24 'data visualization'/exp<br>#25 'computer graphics'/exp<br>#26 'computer graphic*':ti,ab<br>#27 'clinical audit'/exp<br>#28 'medical audit':ab,ti<br>#29 'interactive visualization tool*':ab,ti<br>#30 'feedback':ab,ti<br>#31 'benchmarking':ab,ti<br>#32 ('paperless':ti,ab OR digital:ti,ab OR electronic:ti,ab OR online:ti,ab) AND ('health data':ti,ab OR 'healthcare system*':ti,ab OR 'health care system*':ti,ab OR 'information processing':ti,ab OR |

| | |
|---|---|
| | 'personalised prescription*':ti,ab OR 'personalized prescription*':ti,ab)<br>#33 #13 OR #14 OR #15 OR #16 OR #17 OR #18 OR #19 OR #20 OR #21 OR #22 OR #23 OR #24 OR #25 OR #26 OR #27 OR #28 OR #29 OR #30 OR #31 OR #32<br>#34 'drug prescriptions':ab,ti<br>#35 drug:ab,ti AND prescri*:ab,ti<br>#36 'prescribing error'/exp<br>#37 'inappropriate prescribing':ab,ti<br>#38 ((inappropriate OR over) NEXT/3 prescri*):ab,ti<br>#39 reduc*:ti,ab AND prescription*:ti,ab<br>#40 #34 OR #35 OR #36 OR #37 OR #38 OR #39<br>#41 #12 AND #33 AND #40<br>#41 NOT [medline]/lim |
| Number of references found | 1664 |

| Electronic search report No. 4 | |
|---|---|
| Electronic database | CINAHL Plus with Full Text |
| Platform | EBSCOhost |
| Date of search | 17th November 2023 |
| Range of date | None |
| Restriction of language | None |
| Other limits | None |
| Search Strategy (results) | S1 TI "general practitioner" OR AB "general practitioner"<br>S2 TI "general practice physician*" OR AB "general practice physician*"<br>S3 TI GPs OR AB GPs<br>S4 TI ( ("Family Practice" or "general practice") ) OR AB ( ("Family Practice" or "general practice") )<br>S5 TI ( (family and (practice* or doctor*)) ) OR AB ( (family and (practice* or doctor*)) )<br>S6 TI ( (Clinician* or physician*) ) OR AB ( (Clinician* or physician*) )<br>S7 TI "primary care practice*" OR AB "primary care practice*"<br>S8 TI "Primary Health Care" OR AB "Primary Health Care"<br>S9 TI "primary care" OR AB "primary care"<br>S10 S1 OR S2 OR S3 OR S4 OR S5 OR S6 OR S7 OR S8 OR S9<br>S11 TI "electronic health record*" OR AB "electronic health record*"<br>S12 TI ( ((electronic or computeri?ed) and (health or medical) and record*) ) OR ( ((electronic or computeri?ed) and (health or medical) and record*) )<br>S13 TI ( ("User-Computer Interface*" or "computer interface*") ) OR AB ( ("User-Computer Interface*" or "computer interface*") )<br>S14 TI ( dashboard or "data visuali?ation" ) OR AB ( dashboard or "data visuali*ation" )<br>S15 TI "dashboard" OR AB "dashboard"<br>S16 TI "visual analytic*" OR AB "visual analytic*" |

| | |
|---|---|
| | S17 TI ( ("Health Information System*" OR "medical information system*") ) OR AB ( ("Health Information System*" or "medical information system*") )<br>S18 TI "Health Information System* Data" OR AB "Health Information System* Data"<br>S19 TI "Virtual Desktop Infrastructure" OR AB "Virtual Desktop Infrastructure"<br>S20 TI "data visualisation*" OR AB "data visualisation*"<br>S21 TI "computer graphic*" OR AB "computer graphic*"<br>S22 TI "visual health record" OR AB "visual health record"<br>S23 TI "Interactive visualization tool*" OR AB "Interactive visualization tool*"<br>S24 TI "clinical audit" OR AB "clinical audit"<br>S25 TI "medical audit*" OR AB "medical audit*"<br>S26 TI "feedback" OR AB "feedback"<br>S27 TI "benchmarking" OR AB "benchmarking"<br>S28 (TI "paperless" AB "paperless" OR TI "digital" OR AB "digital" OR TI "electronic OR AB "Digital" OR TI "Online" or AB "Online") AND ( TI "health data" OR AB "health data" OR TI "healthcare system#" OR AB "healthcare system#" OR TI "information processing" OR AB ""information processing" OR TI "personalised prescription#" OR AB "personalised prescription#" OR TI "personalized prescription#" OR AB personalized prescription#")<br>S29 S11 OR S12 OR S13 OR S14 OR S15 OR S16 OR S17 OR S18 OR S19 OR S20 OR S21 OR S22 OR S23 OR S24 OR S25 OR S26 or S27 OR S28<br>S30 TI "drug Prescription*" or AB "drug Prescription*"<br>S31 TI ( (Drug AND Prescri*) ) OR AB ( (Drug AND Prescri*) )<br>S32 TI ( ("inappropriate Prescribing" or "prescribing error") ) OR AB ( ("inappropriate Prescribing" or "prescribing error") )<br>S33 TI ( ((Inappropriate or over) and Prescri*) ) OR AB ( ((Inappropriate or over) and Prescri*) )<br>S34 TI reduc# OR AB reduc# AND TI prescription# OR AB prescription#<br>S35 S30 OR S31 OR S32 OR S33 OR S34<br>S35 S10 AND S29 AND S34<br>S36 S10 AND S29 AND S35 |
| Number of references found | 1805 |

| Electronic search report No. 5 | |
|---|---|
| Electronic database | APA PsychInfo |
| Platform | EBSCOhost |
| Date of search | 17th November 2023 |
| Range of date | None |
| Restriction of language | None |
| Other limits | None |
| Search Strategy (results) | S1 TI "general practitioner" OR AB "general practitioner" |

| | |
|---|---|
| | S2 TI "general practice physician*" OR AB "general practice physician*"<br>S3 TI GPs OR AB GPs<br>S4 TI ( ("Family Practice" or "general practice") ) OR AB ( ("Family Practice" or "general practice") )<br>S5 TI ( (family and (practice* or doctor*)) ) OR AB ( (family and (practice* or doctor*)) )<br>S6 TI ( (Clinician* or physician*) ) OR AB ( (Clinician* or physician*) )<br>S7 TI "primary care practice*" OR AB "primary care practice*"<br>S8 TI "Primary Health Care" OR AB "Primary Health Care"<br>S9 TI "primary care" OR AB "primary care"<br>S10 S1 OR S2 OR S3 OR S4 OR S5 OR S6 OR S7 OR S8 OR S9<br>S11 TI "electronic health record*" OR AB "electronic health record*"<br>S12 TI ( ((electronic or computeri?ed) and (health or medical) and record*) ) OR ( ((electronic or computeri?ed) and (health or medical) and record*) )<br>S13 TI ( ("User-Computer Interface*" or "computer interface*") ) OR AB ( ("User-Computer Interface*" or "computer interface*") )<br>S14 TI ( dashboard or "data visuali?ation" ) OR AB ( dashboard or "data visuali*ation" )<br>S15 TI "dashboard" OR AB "dashboard"<br>S16 TI "visual analytic*" OR AB "visual analytic*"<br>S17 TI ( ("Health Information System*" OR "medical information system*") ) OR AB ( ("Health Information System*" or "medical information system*") )<br>S18 TI "Health Information System* Data" OR AB "Health Information System* Data"<br>S19 TI "Virtual Desktop Infrastructure" OR AB "Virtual Desktop Infrastructure"<br>S20 TI "data visualisation*" OR AB "data visualisation*"<br>S21 TI "computer graphic*" OR AB "computer graphic*"<br>S22 TI "visual health record" OR AB "visual health record"<br>S23 TI "Interactive visualization tool*" OR AB "Interactive visualization tool*"<br>S24 TI "clinical audit" OR AB "clinical audit"<br>S25 TI "medical audit*" OR AB "medical audit*"<br>S26 TI "feedback" OR AB "feedback"<br>S27 TI "benchmarking" OR AB "benchmarking"<br>S28 (TI "paperless" AB "paperless" OR TI "digital" OR AB "digital" OR TI "electronic OR AB "Digital" OR TI "Online" or AB "Online") AND ( TI "health data" OR AB "health data" OR TI "healthcare system#" OR AB "healthcare system#" OR TI "information processing" OR AB ""information processing" OR TI "personalised prescription#" OR AB "personalised prescription#" OR TI "personalized prescription#" OR AB personalized prescription#")<br>S29 S11 OR S12 OR S13 OR S14 OR S15 OR S16 OR S17 OR S18 OR S19 OR S20 OR S21 OR S22 OR S23 OR S24 OR S25 OR S26 or S27 OR S28 |

| | |
|---|---|
| | S30 TI "drug Prescription*" or AB "drug Prescription*"<br>S31 TI ( (Drug AND Prescri*) ) OR AB ( (Drug AND Prescri*) )<br>S32 TI ( ("inappropriate Prescribing" or "prescribing error") ) OR AB ( ("inappropriate Prescribing" or "prescribing error") )<br>S33 TI ( ((Inappropriate or over) and Prescri*) ) OR AB ( ((Inappropriate or over) and Prescri*) )<br>S34 TI reduc# OR AB reduc# AND TI prescription# OR AB prescription#<br>S35 S30 OR S31 OR S32 OR S33 OR S34<br>S35 S10 AND S29 AND S34<br>S36 S10 AND S29 AND S35 |
| Number of references found | 581 |

| Electronic search report No. 6 | |
|---|---|
| Electronic database | PubMed |
| Platform | National Library of Medicine |
| Date of search | 22nd November 2023 |
| Range of date | None |
| Restriction of language | None |
| Other limits | None |
| Search Strategy (results) | (((((((((((((((("Primary Health Care"[Mesh]) OR "Physicians, Family"[Mesh]) OR "Family Practice"[Mesh]) OR "General Practitioners"[Mesh]) ) OR ("general practice physician*"[Title/Abstract])) OR ("general practitioner*"[Title/Abstract])) OR ("GPs"[Title/Abstract])) OR ("general practice"[Title/Abstract])) OR ("family practice*"[Title/Abstract])) OR ("family doctor*"[Title/Abstract])) OR (clinican*[Title/Abstract])) OR (physican*[Title/Abstract])) OR ("primary care practice*"[Title/Abstract])) AND ((((((((((((("Drug Prescriptions"[Mesh]) OR "Patient Care"[Mesh]) OR "Inappropriate Prescribing"[Mesh]) OR ("prescribing error"[Title/Abstract])) OR ("drug prescription$"[Title/Abstract:~10])) OR ("drug prescribing" [Title/Abstract:~10])) OR ("inappropriate prescription" [Title/Abstract:~3])) OR ("inappropriate prescriptions" [Title/Abstract:~3])) OR ("inappropriate prescribing" [Title/Abstract:~3])) OR ("over prescription" [Title/Abstract:~3])) OR ("over prescriptions" [Title/Abstract:~3])) OR ("over prescribing" [Title/Abstract:~3])) OR ("user computer"[Title/Abstract])) OR ("user computer" [Title/Abstract:~3]))) AND (((((((((((((((((((((((("Electronic Health Records"[Mesh]) OR "User-Computer Interface"[Mesh]) OR "Health Information Systems"[Mesh]) OR "Computer Graphics"[Mesh]) OR "Clinical Audit"[Mesh]) OR "Medical Audit"[Mesh]) OR ("computer interface"[Title/Abstract])) OR (dashboard*[Title/Abstract])) OR ("visual analytic*"[Title/Abstract])) OR ("medical information system"[Title/Abstract])) OR ("data visualisation"[Title/Abstract])) OR ("data visualization"[Title/Abstract])) OR ("computer |

| | |
|---|---|
| | graphic*"[Title/Abstract])) OR ("visual health record*"[Title/Abstract])) OR ("health information system* data"[Title/Abstract])) OR ("virtual desktop infrastructure"[Title/Abstract])) OR ("interactive visualization tool*"[Title/Abstract])) OR ("user computer"[Title/Abstract])) OR ("user computer" [Title/Abstract:~3])) OR (((((((paperless[Title/Abstract] AND "health data"[Title/Abstract])) OR (paperless[Title/Abstract] AND "healthcare system*"[Title/Abstract])) OR (paperless[Title/Abstract] AND "health care system*"[Title/Abstract])) OR (paperless[Title/Abstract] AND "information processing"[Title/Abstract])) OR (paperless[Title/Abstract] AND "personalised prescription"[Title/Abstract])) OR (paperless[Title/Abstract] AND "personalized prescription"[Title/Abstract]))) OR (((((((digital[Title/Abstract] AND "health data"[Title/Abstract]) OR (digital[Title/Abstract] AND "healthcare system*"[Title/Abstract])) OR (digtial[Title/Abstract] AND "health care system*"[Title/Abstract])) OR (digital[Title/Abstract] AND "information processing"[Title/Abstract])) OR (digital[Title/Abstract] AND "information processing"[Title/Abstract])) OR (digital[Title/Abstract] AND "personalised prescription*"[Title/Abstract])) OR (digital[Title/Abstract] AND "personalized prescription*"[Title/Abstract]))) OR (((((((electronic[Title/Abstract] AND "health data"[Title/Abstract]) OR (electronic[Title/Abstract] AND "healthcare system*"[Title/Abstract])) OR (electronic[Title/Abstract] AND "health care system*"[Title/Abstract])) OR (electronic[Title/Abstract] AND "information processing"[Title/Abstract])) OR (electronic[Title/Abstract] AND "personalised prescription"[Title/Abstract])) OR (electronic[Title/Abstract] AND "personalised prescription*"[Title/Abstract])) OR (online[Title/Abstract] AND "personalized prescription*"[Title/Abstract]))) OR ((((((online[Title/Abstract] AND "health data"[Title/Abstract]) OR (online[Title/Abstract] AND "healthcare system*"[Title/Abstract])) OR (online[Title/Abstract] AND "health care system*"[Title/Abstract])) OR (online[Title/Abstract] AND "information processing"[Title/Abstract])) OR (online[Title/Abstract] AND "personalised prescription*"[Title/Abstract])) OR (online[Title/Abstract] AND "personalized prescription*"[Title/Abstract])) ) OR (benchmarking[Title/Abstract])) OR ("feedback"[Title/Abstract])) OR (reduc*[Title/Abstract] AND prescription[Title/Abstract])) |
| Number of references found | 5185 |

| Electronic search report No. 7 | |
|---|---|
| Electronic database | SCOPUS |
| Platform | SCOPUS |

| | |
|---|---|
| Date of search | 20th November 2023 |
| Range of date | None |
| Restriction of language | None |
| Other limits | None |
| Search Strategy (results) | ( TITLE-ABS ( ( "drug prescriptions" ) OR ( "drug" AND "prescri*" ) OR "inappropriate prescribing" OR "prescribing error*" OR ( ( inappropriate OR over ) AND ( "prescri*" ) ) OR ( "reduc*" AND "prescription*" ) ) OR ( ( "inappropriate" ) AND ( "prescri*" OR prescribing " or " prescribing AND error ")) or ((" over ") and (" prescri* " or prescribing" OR "prescribing error" ) ) ) AND ( TITLE-ABS ( "electronic health record" OR ( ( "electronic" OR "computeriz?ed" ) AND ( "health" OR "medical" ) AND "record*" ) OR "user-computer interface" OR "computer interface" OR ( ( "user-computer" OR "user computer" ) AND "interface*" ) OR "dashboard*" OR "visual analytic*" OR "health information system*" OR "medical information system*" OR "health information system* data" OR "virtual desktop infrastructure" OR "data visualisation*" OR "computer graphic*" OR "clinical audit" OR "medical audit" OR "feedback" OR "benchmarking" OR "visual health record" OR "interactive visualization tool*" OR ( "paperless" OR "digital" OR "electronic" OR "online" ) AND ( "health data" OR "healthcare system*" OR "health care system" OR "information processing" OR "personalised prescription*" OR "personalized prescription*" ) ) ) AND ( TITLE-ABS ( "general practitioner*" OR "general practice physician*" OR "gps" OR "general practice" OR "family practice" OR ( "family" AND ( "practice*" OR "doctor*" ) ) OR "clinician*" OR "physician*" OR "primary care practice*" OR "primary health care" OR "primary care" ) ) |
| Number of references found | 239 |

#### S3 Appendix Study Inclusion Criteria

| | Inclusion criteria | Exclusion criteria |
|---|---|---|
| **Study design** | Interventional design including RCTs, cRCTs, SWRCTs<br>Controlled before and after studies<br>Interrupted time series analyses | Observational design (e.g. cohort studies), systematic reviews, commentaries, editorials |
| **Population** | Generalist primary care prescribers | Secondary care prescribers<br>Primary care prescribers working in a secondary care setting<br>Specialist primary care prescribers working in a specific field (e.g. dentists) |
| **Intervention** | An interactive dashboard designed to provide feedback on prescribing data to prescribers and including the following characteristics:<br>• **Visual display of data**: Data is presented in the form of graphs or tables.<br>**Real-time data**: Offers real-time or relatively contemporaneous data, no older than one year.<br>**Frequent data feedback**: Provides data feedback more than once.<br>**Comparative analysis**: Compares data to peers or set standards.<br>• **Interactivity**: Allows direct manipulation with visual analytical tools or provides multiple parameters from the dataset, accessible online or via email.<br>Multifaceted interventions that contained an interactive dashboard as defined above as part of the intervention were included. | Clinical decision support interventions<br>Audit and feedback interventions that give once off feedback |
| **Comparison** | Usual care<br>Other educational or prescribing initiatives (e.g., simple audit and feedback or prescriber education). | |
| **Outcomes** | Prescribing related outcome measures such as implicit/explicit criteria, high-risk or low-value criteria, prescribing rates. | No prescribing related outcomes measured. |

#### S4 Appendix: List of all data points extracted from included studies

***Study details***

- Study ID
- Title
- DOI
- Year
- Lead author
- Country in which the study conducted
- Study design
- Study design description

***Population***

- Population to which the intervention was targeted
- Population on whom outcomes were measured
- Total number of participants (targeted for intervention)
- Total number of individuals with outcomes assessed

***Intervention details***

- True interactive dashboard
- Interactive dashboard part of a multifaceted intervention
- If yes, other components in intervention
- Intervention name
- Why?
- What (materials and procedures)
- Who provided it?
- How and where?
- When and how much?
- Tailoring
- Modifications
- How well (planned versus actual)
- Control group

***Outcomes***

- Prescribing outcomes
- Other outcomes
- Primary outcome description

***Results***

- Primary outcome table RCT:
    - Intervention Baseline
    - Intervention Follow-up
    - Control Baseline
    - Control Follow-up
    - Difference (OR/mean difference) Baseline
    - Difference (OR/mean difference) Follow-up
- Primary outcome table ITSA:
    - Pre-intervention Value
    - Post-intervention Value
    - Pre-post relative difference Value
- Other outcome description
    - Other outcome:
    - Intervention Baseline
    - Intervention Follow-up

    - Control Baseline
    - Control Follow-up
    - Difference Baseline
    - Difference Follow-up
- Summary of results

***Risk of bias***

- Random sequence generation
- Random sequence generation supporting text
- Allocation concealment
- Allocation concealment supporting text
- Baseline outcomes measurement similar
- Baseline outcomes measurement similar supporting text
- Baseline characteristics similar
- Baseline characteristics similar supporting text
- Incomplete outcome data
- Incomplete outcome data supporting text
- Knowledge of the allocated interventions adequately prevented during the study
- Knowledge of the allocated interventions adequately prevented during the study supporting text
- Protection against contamination
- Protection against contamination supporting text
- Selective outcome reporting
- Selective outcome reporting supporting text
- Other risks of bias
- Other risks of bias supporting text
- Remaining questions for ITSA: Intervention independent of other changes
- Remaining questions for ITSA Intervention independent of other changes  supporting text
- Shape of the intervention effect pre-specified
- Shape of the intervention effect pre-specified supporting text
- Intervention unlikely to affect data collection
- Intervention unlikely to affect data collection supporting text
- Knowledge of the allocated interventions adequately prevented during the study
- Knowledge of the allocated interventions adequately prevented during the study supporting text

#### S5 Appendix: List of all studies excluded from full text review

| Reference | Reason for exclusion |
|---|---|
| Urbiztondo I, et al. Decreasing inappropriate use of antibiotics in primary care in four countries in South America—cluster randomized controlled trial. Antibiotics 2017;6(4):. MDPI AG; 2017. DOI: 10.3390/antibiotics6040038 | Wrong intervention |
| Schwartz, et al. Computer-Assisted Antimicrobial Recommendations for Optimal Therapy: Analysis of Prescribing Errors in an Antimicrobial Stewardship Trial. Infection control and hospital epidemiology 2017;38(7):857-859 United States 2017 DOI: 10.1017/ice.2017.74 | Wrong setting |
| Dutcher L, et al. Improving Outpatient Antibiotic Prescribing for Respiratory Tract Infections in Primary Care: A Stepped-Wedge Cluster Randomized Trial. Clin Infect Dis. 2022 Mar 23;74(6):947-956. doi: 10.1093/cid/ciab602. | Wrong intervention |
| Barnett KN, et al. Effective Feedback to Improve Primary Care Prescribing Safety (EFIPPS) a pragmatic three-arm cluster randomised trial: designing the intervention (ClinicalTrials.gov registration NCT01602705). Implement Sci. 2014 Oct 11;9:133. DOI: 10.1186/s13012-014-0133-9. | Protocol |
| López-Picazo JJ, et al. Uso de tecnologías de la información para mejorar la seguridad de la prescripción en Atención Primaria [Using information technology to improve drug safety in primary care]. Rev Calid Asist. 2010 Jan-Feb;25(1):12-20. Spanish. doi: 10.1016/j.cali.2009.07.008. | Wrong study design |
| McIsaac W, et al. A pragmatic randomized trial of a primary care antimicrobial stewardship intervention in Ontario, Canada. BMC Fam Pract. 2021 Sep 15;22(1):185. doi: 10.1186/s12875-021-01536-3. | Wrong intervention |
| Schapira M, et al. A multifactorial intervention to lower potentially inappropriate medication use in older adults in Argentina. Aging Clin Exp Res. 2021 Dec;33(12):3313-3320. doi: 10.1007/s40520-020-01582-4. | Wrong intervention |
| Price M, et al. Applying STOPP Guidelines in Primary Care Through Electronic Medical Record Decision Support: Randomized Control Trial Highlighting the Importance of Data Quality. JMIR Med Inform. 2017 Jun 15;5(2):e15. doi: 10.2196/medinform.6226. | Wrong intervention |
| Peiris, D. Effect of a multi-faceted quality improvement intervention to improve cardiovascular disease risk identification and management in australian primary health care: The torpedo cluster-randomised trial. Global Heart. 2014;9(1):e28. doi; 10.1016/j.gheart.2014.03.1317 | Wrong intervention |
| Mainous AG 3rd et al. Impact of a clinical decision support system on antibiotic prescribing for acute respiratory infections in primary care: quasi-experimental trial. J Am Med Inform Assoc. 2013 Mar-Apr;20(2):317-24. doi: 10.1136/amiajnl-2011-000701. | Wrong intervention |
| Zhuo C, et al. An antibiotic stewardship programme to reduce inappropriate antibiotic prescribing for acute respiratory infections in rural Chinese primary care facilities: study protocol for a clustered | Wrong intervention |

| | |
|---|---|
| randomised controlled trial. Trials. 2020 May 12;21(1):394. doi: 10.1186/s13063-020-04303-4. | |
| Yang J, et al.. Effects of a feedback intervention on antibiotic prescription control in primary care institutions based on a Health Information System: a cluster randomized cross-over controlled trial. J Glob Antimicrob Resist. 2023 Jun;33:51-60. doi: 10.1016/j.jgar.2023.02.006. | Wrong intervention |
| Dreischulte T, et al. Data driven quality improvement in primary care (DQIP): Protocol of a cluster randomised controlled trial in 2 Scottish Health boards. International Journal of Clinical Pharmacy 2012;34(1):232. doi:10.1007/s11096-011-9602-2 | Wrong intervention |
| Andrade AQ, et al. Implementation and Evaluation of a Digitally Enabled Precision Public Health Intervention to Reduce Inappropriate Gabapentinoid Prescription: Cluster Randomized Controlled Trial. J Med Internet Res. 2022 Jan 10;24(1):e33873. doi: 10.2196/33873. | Wrong intervention |
| Mecca MC, et al. Primary care clinicians' use of deprescribing recommendations: A mixed-methods study. Patient Educ Couns. 2022 Aug;105(8):2715-2720. doi: 10.1016/j.pec.2022.04.013. | Wrong study design |
| Madaras-Kelly KJ, et al. Implementation and outcomes of a clinician-directed intervention to improve antibiotic prescribing for acute respiratory tract infections within the Veterans' Affairs Healthcare System. Infect Control Hosp Epidemiol. 2023 May;44(5):746-754. doi: 10.1017/ice.2022.182. | Wrong intervention |
| Soucy JR, et al. Evaluation of an automated feedback intervention to improve antimicrobial prescribing among primary care physicians (OPEN Stewardship): protocol for an interrupted time-series and usability analysis in Ontario, Canada and Southern Israel. BMJ Open. 2021 Jan 13;11(1):e039810. doi: 10.1136/bmjopen-2020-039810. | Protocol |
| Tamblyn R, et al. The medical office of the 21st century (MOXXI): effectiveness of computerized decision-making support in reducing inappropriate prescribing in primary care. CMAJ. 2003 Sep 16;169(6):549-56. | Wrong intervention |
| Vellinga A, et al. Intervention to improve the quality of antimicrobial prescribing for urinary tract infection: a cluster randomized trial. CMAJ. 2016 Feb 2;188(2):108-115. doi: 10.1503/cmaj.150601. | Wrong intervention |
| Vervloet M, et al. Reducing antibiotic prescriptions for respiratory tract infections in family practice: results of a cluster randomized controlled trial evaluating a multifaceted peer-group-based intervention. NPJ Prim Care Respir Med. 2016 Feb 4;26:15083. doi: 10.1038/npjpcrm.2015.83. | Wrong intervention |
| Gentry E, et al. A 20/20 vision: Successful integration of a prescribing dashboard for outpatient antimicrobial stewardship to target 20% reduction by the year 2020. Open Forum Infectious Diseases 2019;6():S49. doi:10.1093/ofid/ofz359.109. | Wrong setting |
| Gjelstad S, et al. Improving antibiotic prescribing in acute respiratory tract infections: cluster randomised trial from Norwegian general practice (prescription peer academic detailing (Rx-PAD) study). BMJ. 2013 Jul 26;347:f4403. doi: 10.1136/bmj.f4403. | Wrong intervention |
| Glinz D, et al. Antibiotic prescription monitoring and feedback in primary care in Switzerland: Design and rationale of a nationwide | Protocol |

| | |
|---|---|
| pragmatic randomized controlled trial. Contemp Clin Trials Commun. 2021 Jan 20;21:100712. doi: 10.1016/j.conctc.2021.100712. | |
| Hesse U, et al. A new tool to evaluate and promote rational use of pharmacotherapy available to all Danish general practitioners. Pharmacoepidemiology and Drug Safety 2019;28():536. Doi:10.1002/pds.4864. | Wrong study design |
| Greiver M, et al. Improving care for elderly patients living with polypharmacy: protocol for a pragmatic cluster randomized trial in community-based primary care practices in Canada. Implement Sci. 2019 Jun 6;14(1):55. doi: 10.1186/s13012-019-0904-4. | Protocol |
| MacBride-Stewart S, et al. Evaluation of a complex intervention to improve primary care prescribing: a phase IV segmented regression interrupted time series analysis. Br J Gen Pract. 2017 May;67(658):e352-e360. doi: 10.3399/bjgp17X690437. | Wrong intervention |
| Seume P, et al. Protocol for an 'efficient design' cluster randomised controlled trial to evaluate a complex intervention to improve antibiotic prescribing for CHIldren presenting to primary care with acute COugh and respiratory tract infection: the CHICO study. BMJ Open. 2021 Mar 29;11(3):e041769. doi: 10.1136/bmjopen-2020-041769. | Wrong intervention |
| Chang Y, et al. Changing antibiotic prescribing practices in outpatient primary care settings in China: Study protocol for a health information system-based cluster-randomised crossover controlled trial. PLoS One. 2022 Jan 7;17(1):e0259065. doi: 10.1371/journal.pone.0259065. | Wrong intervention |
| Brown T, et al. Reducing high-risk geriatric polypharmacy via electronic health record nudges. Journal of General Internal Medicine. 2020;35(SUPPL1):S253. doi:10.1007/s11606-020-05890-3. | Wrong intervention |
| Fàbregas M, et al. Effectiveness of an intervention designed to optimize statins use: a primary prevention randomized clinical trial. BMC Fam Pract. 2014 Jul 15;15:135. doi: 10.1186/1471-2296-15-135. | Protocol |
| Fernández-Urrusuno R, et al. Successful improvement of antibiotic prescribing at Primary Care in Andalusia following the implementation of an antimicrobial guide through multifaceted interventions: An interrupted time-series analysis. PLoS One. 2020 May 15;15(5):e0233062. doi: 10.1371/journal.pone.0233062. | Wrong intervention |
| Optimizing Electronic Health Record Prompts With Behavioral Economics to Improve Prescribing for Older Adults, https://www.nia.nih.gov/research/dbsr/workshops/optimizing-older-adult-care-through-use-electronic-health-records | Wrong study design |
| Impact of medAL-mentor, a Real-time Mentoring and Benchmarking Tool, on Antibiotic Prescription Among Children in Primary Health Care Facilities in Tanzania, https://clinicaltrials.gov/study/NCT05901155 | Wrong intervention |
| Tamblyn R, et al. The effectiveness of a new generation of computerized drug alerts in reducing the risk of injury from drug side effects: a cluster randomized trial. J Am Med Inform Assoc. 2012 Jul-Aug;19(4):635-43. doi: 10.1136/amiajnl-2011-000609. | Wrong intervention |
| Fried TR, et al. Effect of the Tool to Reduce Inappropriate Medications on Medication Communication and Deprescribing. J Am Geriatr Soc. | Wrong intervention |

| | |
|---|---|
| 2017 Oct;65(10):2265-2271. doi: 10.1111/jgs.15042. Epub 2017 Aug 14. | |
| Adusumalli S, et al. Effect of Nudges to Clinicians, Patients, or Both to Increase Statin Prescribing: A Cluster Randomized Clinical Trial. JAMA Cardiol. 2023 Jan 1;8(1):23-30. doi: 10.1001/jamacardio.2022.4373. | Wrong intervention |
| Peiris D, et al. The Treatment of cardiovascular Risk in Primary care using Electronic Decision supOrt (TORPEDO) study-intervention development and protocol for a cluster randomised, controlled trial of an electronic decision support and quality improvement intervention in Australian primary healthcare. BMJ Open. 2012 Nov 19;2(6):e002177. doi: 10.1136/bmjopen-2012-002177. | Protocol |
| B Sussman J, et al. Quality Improvement and Personalization for Statins: the QUIPS Quality Improvement Randomized Trial of Veterans' Primary Care Statin Use. J Gen Intern Med. 2018 Dec;33(12):2132-2137. doi: 10.1007/s11606-018-4681-6. | Wrong intervention |
| Zhang Z, et al. Cost-effectiveness analysis of a multi-dimensional intervention to reduce inappropriate antibiotic prescribing for children with upper respiratory tract infections in China. Trop Med Int Health. 2018 Oct;23(10):1092-1100. doi: 10.1111/tmi.13132. | Wrong study design |
| Vanstone JR, et al. Using audit and feedback to encourage primary healthcare prescribers to record indications for antimicrobial prescriptions: a quality improvement initiative. BMJ Open Qual. 2022 Mar;11(1):e001760. doi: 10.1136/bmjoq-2021-001760. | Wrong outcomes |
| Murray MD, et al. Failure of computerized treatment suggestions to improve health outcomes of outpatients with uncomplicated hypertension: results of a randomized controlled trial. Pharmacotherapy. 2004 Mar;24(3):324-37. doi: 10.1592/phco.24.4.324.33173. | Wrong intervention |
| Neprash HT, et al. Effect of Integrating Access to a Prescription Drug Monitoring Program Within the Electronic Health Record on the Frequency of Queries by Primary Care Clinicians: A Cluster Randomized Clinical Trial. JAMA Health Forum. 2022 Jun 5;3(6):e221852. doi: 10.1001/jamahealthforum.2022.1852. | Wrong intervention |
| Milani RV, et al. Reducing inappropriate outpatient antibiotic prescribing: normative comparison using unblinded provider reports. BMJ Open Qual. 2019 Feb 13;8(1):e000351. doi: 10.1136/bmjoq-2018-000351. | Wrong intervention |
| Herbert CP, et al. Better Prescribing Project: a randomized controlled trial of the impact of case-based educational modules and personal prescribing feedback on prescribing for hypertension in primary care. Fam Pract. 2004 Oct;21(5):575-81. doi: 10.1093/fampra/cmh515. | Wrong intervention |
| Rodgers S, et al. Scaling-up a pharmacist-led information technology intervention (PINCER) to reduce hazardous prescribing in general practices: Multiple interrupted time series study. PLoS Med. 2022 Nov 16;19(11):e1004133. doi: 10.1371/journal.pmed.1004133. | Wrong intervention |
| Dhingra L, et al. Pain Management in Primary Care: A Randomized Controlled Trial of a Computerized Decision Support Tool. Am J Med. 2021 Dec;134(12):1546-1554. doi: 10.1016/j.amjmed.2021.07.014. | Wrong intervention |

| | |
|---|---|
| O'Connor PJ, et al. Simulated physician learning intervention to improve safety and quality of diabetes care: a randomized trial. Diabetes Care. 2009 Apr;32(4):585-90. doi: 10.2337/dc08-0944. | Wrong intervention |
| Mombelli M, et al. Antimicrobial stewardship en pratique communautaire [Antimicrobial stewardship in primary care setting]. Rev Med Suisse. 2016 Apr 13;12(514):744-8. French. | Wrong study design |
| Buehrle DJ, et al. Sustained Reductions in Overall and Unnecessary Antibiotic Prescribing at Primary Care Clinics in a Veterans Affairs Healthcare System Following a Multifaceted Stewardship Intervention. Clin Infect Dis. 2020 Nov 5;71(8):e316-e322. doi: 10.1093/cid/ciz1180. | Wrong study design |
| Parveen S, et al.. Collaboration to reduce antibiotic use and resistance and identify opportunities for improvement and awareness (CARA). Rural Remote Health. 2023 Jan;23(1):8153. doi: 10.22605/RRH8153. | Abstract for oral presentation |
| Rogero-Blanco E, et al. Use of an Electronic Clinical Decision Support System in Primary Care to Assess Inappropriate Polypharmacy in Young Seniors With Multimorbidity: Observational, Descriptive, Cross-Sectional Study. JMIR Med Inform. 2020 Mar 3;8(3):e14130. doi: 10.2196/14130. Erratum in: JMIR Med Inform. 2020 Nov 19;8(11):e25678. doi: 10.2196/25678. | Wrong study design |
| Soames J, et al. Reducing unnecessary antibiotic prescribing in primary care: A point of-care electronic intervention using electronic healthcare records (reduce trial) | Abstract for oral presentation |
| Clyne B, et al. Effectiveness of medicines review with web-based pharmaceutical treatment algorithms in reducing potentially inappropriate prescribing in older people in primary care: a cluster randomized trial (OPTI-SCRIPT study protocol). Trials. 2013 Mar 13;14:72. Doi: 10.1186/1745-6215-14-72. | Wrong intervention |
| Gulliford MC, et al. Effectiveness and safety of electronically delivered prescribing feedback and decision support on antibiotic use for respiratory illness in primary care: REDUCE cluster randomised trial. BMJ. 2019 Feb 12;364:l236. Doi: 10.1136/bmj.l236. | Wrong intervention |
| Joshi S. Feedback to clinicians on antibiotic prescription habits: how effective are they? Indian J Med Microbiol. 2015 Apr-Jun;33(2):260-1. Doi: 10.4103/0255-0857.154868. | Wrong study design |
| Persell SD, et al. Changes in performance after implementation of a multifaceted electronic-health-record-based quality improvement system. Med Care. 2011 Feb;49(2):117-25. Doi: 10.1097/MLR.0b013e318202913d. | Wrong outcomes |
| Persell SD, et al. Behavioral economics-informed interventions to reduce inappropriate antibiotic prescribing: A pilot cluster randomized trial. Journal of General Internal Medicine. 2014;29():S40-S41. | Wrong intervention |
| Liebschutz JM, et al. Improving Adherence to Long-term Opioid Therapy Guidelines to Reduce Opioid Misuse in Primary Care: A Cluster-Randomized Clinical Trial. JAMA Intern Med. 2017 Sep 1;177(9):1265-1272. doi: 10.1001/jamainternmed.2017.2468. | Wrong intervention |
| Katz SE, et al. Improvements in appropriate ambulatory antibiotic prescribing using a bundled antibiotic stewardship intervention in general pediatrics practices. Infect Control Hosp Epidemiol. 2022 Dec;43(12):1894-1900. doi: 10.1017/ice.2021.534. | Wrong study design |

| | |
|---|---|
| Tierney WM, et al. Can computer-generated evidence-based care suggestions enhance evidence-based management of asthma and chronic obstructive pulmonary disease? A randomized, controlled trial. Health Serv Res. 2005 Apr;40(2):477-97. doi: 10.1111/j.1475-6773.2005.00368.x. | Wrong intervention |
| Payne RA, et al. Improving Medicines use in People with Polypharmacy in Primary Care (IMPPP): Protocol for a multicentre cluster randomised trial comparing a complex intervention for medication optimization against usual care. NIHR Open Res. 2022 Nov 8;2:54. doi: 10.3310/nihropenres.13285.1. | Protocol |
| Campbell NL, Holden RJ, Tang Q, Boustani MA, Teal E, Hillstrom J, Tu W, Clark DO, Callahan CM. Multicomponent behavioral intervention to reduce exposure to anticholinergics in primary care older adults. J Am Geriatr Soc. 2021 Jun;69(6):1490-1499. doi: 10.1111/jgs.17121. | Wrong intervention |
| Spiegel BMR, et al. Cluster-Randomized Comparative Effectiveness Trial of Physician-Directed Clinical Decision Support Versus Patient-Directed Education to Promote Appropriate Use of Opioids for Chronic Pain. J Pain. 2023 Oct;24(10):1745-1758. doi: 10.1016/j.jpain.2023.06.001. | Wrong intervention |
| Shuldiner J, et al. Implementation Laboratory study team. Optimizing responsiveness to feedback about antibiotic prescribing in primary care: protocol for two interrelated randomized implementation trials with embedded process evaluations. Implement Sci. 2022 Feb 14;17(1):17. doi: 10.1186/s13012-022-01194-8. | Wrong intervention |
| Smith S, et al. A standardized methodology for the surveillance of antimicrobial prescribing linked to clinical indications in primary care. J Public Health (Oxf). 2018 Sep 1;40(3):630-638. doi: 10.1093/pubmed/fdx114. | Wrong study design |
| Schaefer K, Hansen AO, Maerkedahl H, Rehfeld C, Birk HO, Henriksen LO. Påvirkning af praktiserende laegers ordinationer via kliniske retningslinjer og feedback--sekundaerpublikation [Influencing the prescriptions of general practitioners via guidelines and feedback--secondary publication]. Ugeskr Laeger. 2008 Dec 1;170(49):4030-2. Danish. | No full text |
| Automated feedback reduces high-risk prescribing in primary care, Drugs and therapeutics bulletin | Wrong study design |
| Hingorani R, et al. Improving antibiotic adherence in treatment of acute upper respiratory infections: a quality improvement process. J Community Hosp Intern Med Perspect. 2015 Jun 15;5(3):27472. doi: 10.3402/jchimp.v5.27472. | Wrong study design |
| Dreischulte T, et al. A cluster randomised stepped wedge trial to evaluate the effectiveness of a multifaceted information technology-based intervention in reducing high-risk prescribing of non-steroidal anti-inflammatory drugs and antiplatelets in primary medical care: the DQIP study protocol. Implement Sci. 2012 Mar 23;7:24. doi: 10.1186/1748-5908-7-24. | Protocol |
| Funaro JR, et al. Impact of Education and Data Feedback on Guideline-Concordant Prescribing for Urinary Tract Infections in the Outpatient Setting. Open Forum Infect Dis. 2021 Apr 28;9(3):ofab214. doi: 10.1093/ofid/ofab214. | Wrong study design |

| | |
|---|---|
| Lauffenburger JC, et al. Rationale and design of the Novel Uses of adaptive Designs to Guide provider Engagement in Electronic Health Records (NUDGE-EHR) pragmatic adaptive randomized trial: a trial protocol. Implement Sci. 2021 Jan 7;16(1):9. doi: 10.1186/s13012-020-01078-9. | Wrong intervention |
| Martens JD, et al. Design and evaluation of a computer reminder system to improve prescribing behaviour of GPs. Stud Health Technol Inform. 2006;124:617-23. | Wrong intervention |
| Peiris D, et al. Effect of a computer-guided, quality improvement program for cardiovascular disease risk management in primary health care: the treatment of cardiovascular risk using electronic decision support cluster-randomized trial. Circ Cardiovasc Qual Outcomes. 2015 Jan;8(1):87-95. doi: 10.1161/CIRCOUTCOMES.114.001235. Epub 2015 Jan 13. Erratum in: Circ Cardiovasc Qual Outcomes. 2018 Sep;11(9):e000049. doi: 10.1161/HCQ.0000000000000049. | Wrong intervention |
| Hernandez-Santiago V, et al. Time series analysis of the impact of an intervention in Tayside, Scotland to reduce primary care broad-spectrum antimicrobial use. J Antimicrob Chemother. 2015 Aug;70(8):2397-404. doi: 10.1093/jac/dkv095. | Wrong study design |
| Yang J, et al. Effects of a feedback intervention on antibiotic prescription control in primary care institutions based on a Health Information System: a cluster randomized cross-over controlled trial. J Glob Antimicrob Resist. 2023 Jun;33:51-60. doi: 10.1016/j.jgar.2023.02.006. | Wrong intervention |
| Barnett KN, et al. Effective Feedback to Improve Primary Care Prescribing Safety (EFIPPS) a pragmatic three-arm cluster randomised trial: designing the intervention (ClinicalTrials.gov registration NCT01602705). Implement Sci. 2014 Oct 11;9:133. doi: 10.1186/s13012-014-0133-9. | Protocol |
| Chang Y, et al. Effect of a computer network-based feedback program on antibiotic prescription rates of primary care physicians: A cluster randomized crossover-controlled trial. J Infect Public Health. 2020 Sep;13(9):1297-1303. doi: 10.1016/j.jiph.2020.05.027. | Wrong intervention |
| Gulliford MC, et al. Electronic health records for intervention research: a cluster randomized trial to reduce antibiotic prescribing in primary care (eCRT study). Ann Fam Med. 2014 Jul;12(4):344-51. doi: 10.1370/afm.1659. | Wrong intervention |
| Guthrie B,et al. Protocol for the Effective Feedback to Improve Primary Care Prescribing Safety (EFIPPS) study: a cluster randomised controlled trial using ePrescribing data. BMJ Open. 2012 Dec 13;2(6):e002359. doi: 10.1136/bmjopen-2012-002359. | Protocol |
| Harrigan JJ, et al. Antibiotic Prescribing Patterns for Respiratory Tract Illnesses Following the Conclusion of an Education and Feedback Intervention in Primary Care. Clin Infect Dis. 2024 May 15;78(5):1120-1127. doi: 10.1093/cid/ciad754. | Abstract for oral presentation |
| Juszczyk D, et al. Electronically delivered, multicomponent intervention to reduce unnecessary antibiotic prescribing for respiratory infections in primary care: a cluster randomised trial using | Protocol |

| | |
|---|---|
| electronic health records-REDUCE Trial study original protocol. BMJ Open. 2016 Aug 4;6(8):e010892. doi: 10.1136/bmjopen-2015-010892. | |
| Litvin CB, et al. Use of an electronic health record clinical decision support tool to improve antibiotic prescribing for acute respiratory infections: the ABX-TRIP study. J Gen Intern Med. 2013 Jun;28(6):810-6. doi: 10.1007/s11606-012-2267-2. | Wrong study design |
| Gonzales R, et al. A cluster randomized trial of decision support strategies for reducing antibiotic use in acute bronchitis. JAMA Intern Med. 2013 Feb 25;173(4):267-73. doi: 10.1001/jamainternmed.2013.1589. | Wrong intervention |
| Reducing Cardiovascular disease (CVD): Translating an evidence based quality improvement tool into 'real-world' general practice, ANZCTR registration: ACTRN 12615000108516 | Wrong intervention |
| Avent ML, et al. Reducing antibiotic prescribing in general practice in Australia: a cluster randomised controlled trial of a multimodal intervention. Aust J Prim Health. 2024 Feb;30(1):NULL. doi: 10.1071/PY23024. | Wrong intervention |
| Tamblyn R, et al. Increasing the detection and response to adherence problems with cardiovascular medication in primary care through computerized drug management systems: a randomized controlled trial. Med Decis Making. 2010 Mar-Apr;30(2):176-88. doi: 10.1177/0272989X09342752. | Wrong intervention |
| The effect of Information Technology in Reducing Potential Drug-drug Interactions in Physicians' Prescriptions, https://irct.behdasht.gov.ir/trial/17021 | Protocol |
| Hemkens LG, et al. Personalized prescription feedback to reduce antibiotic overuse in primary care: rationale and design of a nationwide pragmatic randomized trial. BMC Infect Dis. 2016 Aug 17;16:421. doi: 10.1186/s12879-016-1739-0. | Protocol |
| Testing CDS in OSCAR EMR Using STOPP Criteria, https://trialsearch.who.int/Trial2.aspx?TrialID=NCT02130895 | Wrong intervention |
| Bonney A, et al. Randomised trial of general practitioner online education for prescribing and test ordering. BMJ Open Qual. 2023 Oct;12(4):e002351. doi: 10.1136/bmjoq-2023-002351. | Wrong intervention |
| Wessell AM, et al. Medication Safety in Primary Care Practice: results from a PPRNet quality improvement intervention. Am J Med Qual. 2013 Jan-Feb;28(1):16-24. doi: 10.1177/1062860612445070. | Wrong study design |
| Müller BS, et al. Effectiveness of the application of an electronic medication management support system in patients with polypharmacy in general practice: a study protocol of cluster-randomised controlled trial (AdAM). BMJ Open. 2021 Sep 28;11(9):e048191. doi: 10.1136/bmjopen-2020-048191. | Wrong intervention |
| Niehoff KM, et al. Development of the Tool to Reduce Inappropriate Medications (TRIM): A Clinical Decision Support System to Improve Medication Prescribing for Older Adults. Pharmacotherapy. 2016 Jun;36(6):694-701. doi: 10.1002/phar.1751. | Wrong intervention |
| Palen TE, et al. Evaluation of laboratory monitoring alerts within a computerized physician order entry system for medication orders. Am J Manag Care. 2006 Jul;12(7):389-95. | Wrong intervention |

| | |
|---|---|
| Høye S, et al. Effects on antibiotic dispensing rates of interventions to promote delayed prescribing for respiratory tract infections in primary care. Br J Gen Pract. 2013 Nov;63(616):e777-86. doi: 10.3399/bjgp13X674468. | Wrong intervention |
| Bonney A, et al. Clinical and healthcare improvement through My Health Record usage and education in general practice (CHIME-GP): a study protocol for a cluster-randomised controlled trial. Trials. 2021 Aug 28;22(1):569. doi: 10.1186/s13063-021-05438-8. | Protocol |
| Awdishu L, et al. The impact of real-time alerting on appropriate prescribing in kidney disease: a cluster randomized controlled trial. J Am Med Inform Assoc. 2016 May;23(3):609-16. doi: 10.1093/jamia/ocv159. | Wrong intervention |
| Persell SD, et al. Development of High-Risk Geriatric Polypharmacy Electronic Clinical Quality Measures and a Pilot Test of EHR Nudges Based on These Measures. J Gen Intern Med. 2022 Aug;37(11):2777-2785. doi: 10.1007/s11606-021-07296-1. | Wrong intervention |
| Schmiemann G, et al. Effects of a multimodal intervention in primary care to reduce second line antibiotic prescriptions for urinary tract infections in women: parallel, cluster randomised, controlled trial. BMJ. 2023 Nov 2;383:e076305. doi: 10.1136/bmj-2023-076305. | Wrong intervention |
| Bregnhøj L, et al. Combined intervention programme reduces inappropriate prescribing in elderly patients exposed to polypharmacy in primary care. Eur J Clin Pharmacol. 2009 Feb;65(2):199-207. doi: 10.1007/s00228-008-0558-7. | Wrong intervention |
| Shively NR, et al. Improved Antibiotic Prescribing within a Veterans Affairs Primary Care System through a Multifaceted Intervention Centered on Peer Comparison of Overall Antibiotic Prescribing Rates. Antimicrob Agents Chemother. 2019 Dec 20;64(1):e00928-19. doi: 10.1128/AAC.00928-19. | Wrong intervention |
| Patel MS, et al. Effect of an Automated Patient Dashboard Using Active Choice and Peer Comparison Performance Feedback to Physicians on Statin Prescribing: The PRESCRIBE Cluster Randomized Clinical Trial. JAMA Netw Open. 2018 Jul 6;1(3):e180818. doi: 10.1001/jamanetworkopen.2018.0818. | Wrong intervention |
| Computer-Based Decision Support in Managing Asthma in Primary Care, https://clinicaltrials.gov/study/NCT00170248 | Wrong intervention |
| Galimberti F, et al. Evaluation of Factors Associated With Appropriate Drug Prescription and Effectiveness of Informative and Educational Interventions-The EDU.RE.DRUG Project. Front Pharmacol. 2022 Apr 25;13:832169. doi: 10.3389/fphar.2022.832169. | Wrong intervention |
| Assessing the impact of information technology-based interventions on the amount of antibiotics prescribed by physicians, https://trialsearch.who.int/Trial2.aspx?TrialID=IRCT2015030815243N2 | Protocol |
| Improving Safety After Hospitalization in Older Persons on High-Risk Medications, https://trialsearch.who.int/Trial2.aspx?TrialID=NCT02781662 | Wrong intervention |
| Albaina O, et al. Assessment of ICT solutions to promote the management of polypharmacy in elderly people with multi-chronic...21st International Conference on Integrated Care (Virtual), | Wrong study design |

| | |
|---|---|
| May 1-31, 2021. International Journal of Integrated Care (IJIC) 2022;22():1-2. doi:10.5334/ijic.ICIC21052. | |
| Cardwell K, et al. General Practice Pharmacist (GPP) Study Group. Supporting prescribing in Irish primary care: protocol for a non-randomised pilot study of a general practice pharmacist (GPP) intervention to optimise prescribing in primary care. Pilot Feasibility Stud. 2018 Jul 5;4:122. doi: 10.1186/s40814-018-0311-7. | Wrong intervention |
| Bourgeois FC, et al.. Impact of a computerized template on antibiotic prescribing for acute respiratory infections in children and adolescents. Clin Pediatr (Phila). 2010 Oct;49(10):976-83. doi: 10.1177/0009922810373649. | Wrong intervention |

### S6 Appendix Characteristics of Included Studies

| Lead author, year, country | Design | Physicians/practices targeted for intervention | Patients outcome assessed | Prescribing outcome |
|---|---|---|---|---|
| Aghlmandi (2023, Switzerland)[1] | RCT | 3426 top antibiotic prescribing physicians | N/A | Antibiotic prescribing rate |
| Curtis (2021, UK)[2] | 3-arm cRCT | 1401 top antibiotic prescribing practices | N/A | Board spectrum antibiotic use |
| Davidson (2023, USA)[3] | ITS | 162 practices | N/A | Antibiotic prescribing rate |
| deLusigan (2021, UK)[4] | CBA | 12 practices | 807<br>8 | Inappropriate aspirin<br>Inappropriate metformin |
| Dutcher (2021, USA)[5] | SW-RCT | 31 practices | N/A | Antibiotic prescribing rate |
| Guthrie (2016, UK)[6] | 3-arm cRCT | 262 practices | 170,659 | Composite measure of high-risk prescribing |
| Hemkins (2017, Switzerland)[7] | RCT | 2900 top abx prescribing physicians | N/A | Antibiotic prescribing rate |
| MacBride-Stewart (2022, UK)[8] | cRCT | 235 practices | N/A | Inappropriate SABA and LABA prescriptions |
| Peek (2020, UK)[9] | ITS | 43 practices | 54,044 | Composite measure of high-risk prescribing |
| Soucy (2024, Israel/ Canada)[10] | ITS | 43 physicians | N/A | Antibiotic prescribing rate |
| Willis (2020, UK)[11]* | cRCT | 40 practices | 34,822 | Inappropriate aspirin or NSAID prescribing |
| Willis (2020, UK)[11]* | cRCT | 32 practices | 48,848 | Inappropriate omission of anticoagulant |

* This study recruited for two trials concurrently; 80 practices were randomised to a trial exploring the effect of feedback of high risk NSAID and antiplatelet prescribing and 64 to explore the effect of feedback on appropriate anticoagulant prescribing for atrial fibrillation.

*Abbreviations: RCT; randomised controlled trial,, cRCT; cluster randomised controlled trial, ITS; interrupted time series, CBA; controlled before and after study, SW-RCT; stepped wedge randomised controlled trial, SABA; short acting beta agonist inhaler, LABA; long acting beta agonist inhaler, NSAID; non-steroidal anti-inflammatory drug.*

References

1. Aghlmandi S, Halbeisen FS, Saccilotto R, et al. Effect of Antibiotic Prescription Audit and Feedback on Antibiotic Prescribing in Primary Care: A Randomized Clinical Trial. JAMA internal medicine. 2023;183(3):213-220.
2. Curtis HJ, Bacon S, Croker R, et al. Evaluating the impact of a very low-cost intervention to increase practices' engagement with data and change prescribing behaviour: a randomized trial in English primary care. Fam Pract. 2021;38(4):373-380.
3. Davidson LE, Gentry EM, Priem JS, Kowalkowski M, Spencer MD. A multimodal intervention to decrease inappropriate outpatient antibiotic prescribing for upper respiratory tract infections in a large integrated healthcare system. Infect Control Hosp Epidemiol. 2023;44(3):392-399.
4. de Lusignan S, Hinton W, Seidu S, et al. Dashboards to reduce inappropriate prescribing of metformin and aspirin: A quality assurance programme in a primary care sentinel network. Prim Care Diabetes. 2021;15(6):1075-1079.
5. Dutcher L, Degnan KO, Adu-Gyamfi AB, et al. Improving Outpatient Antibiotic Prescribing for Respiratory Tract Infections in Primary Care; a Stepped-Wedge Cluster Randomized Trial. Clinical infectious diseases : an official publication of the Infectious Diseases Society of America. 2021;74(6):947-956.
6. Guthrie B, Kavanagh K, Robertson C, et al. Data feedback and behavioural change intervention to improve primary care prescribing safety (EFIPPS): multicentre, three arm, cluster randomised controlled trial. BMJ (Clinical research ed). 2016;354:i4079.
7. Hemkens LG, Saccilotto R, Leon Reyes S, et al. Personalized Prescription Feedback Using Routinely Collected Data to Reduce Antibiotic Use in Primary Care: A Randomized Clinical Trial. JAMA internal medicine. 2017;177(2):176-183.
8. MacBride-Stewart S, Marwick C, Ryan M, Guthrie B. Feedback of actionable individual patient prescription data to improve asthma prescribing: pragmatic cluster randomised trial in 233 UK general practices. Br J Gen Pract. 2022;72(722):e627-633.
9. Peek N, Gude WT, Keers RN, et al. Evaluation of a pharmacist-led actionable audit and feedback intervention for improving medication safety in UK primary care: An interrupted time series analysis. PLoS Med. 2020;17(10):1-17.
10. Soucy Jean-Paul R, Low M, Acharya Kamal R, et al. Evaluation of an automated feedback intervention to improve antibiotic prescribing among primary care physicians (OPEN Stewardship): a multinational controlled interrupted time-series study. Microbiology spectrum. 2024;12(4):e00017-00024.
11. Willis TA, Collinson M, Glidewell L, et al. An adaptable implementation package targeting evidence-based indicators in primary care: A pragmatic cluster-randomised evaluation. PLoS Med. 2020;17(2):1-20.

#### S7 Appendix: Risk of bias graphs

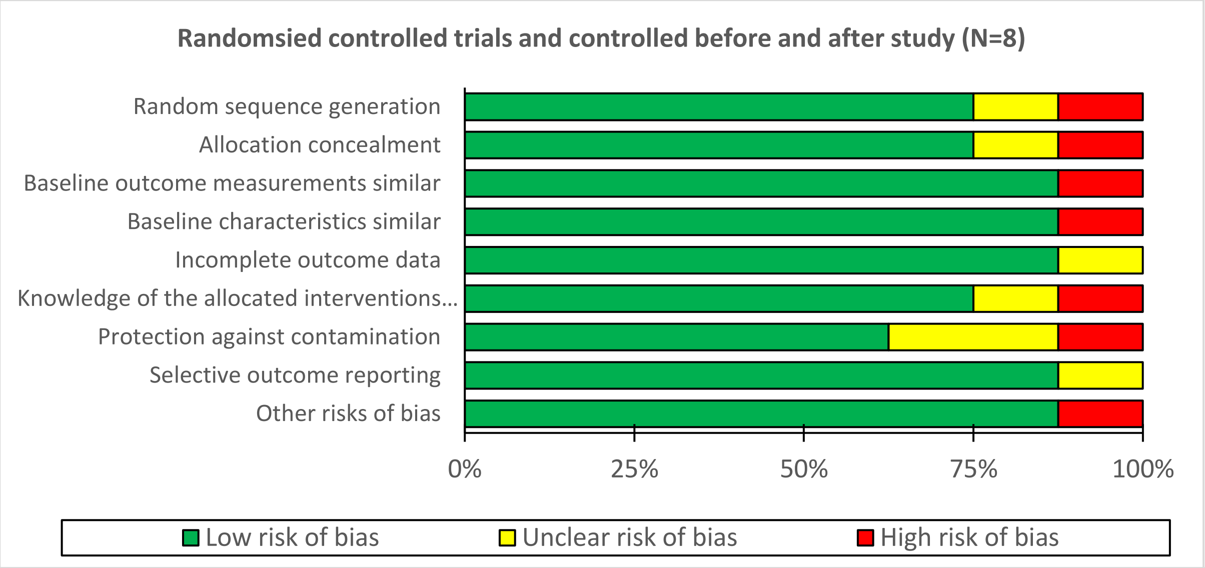

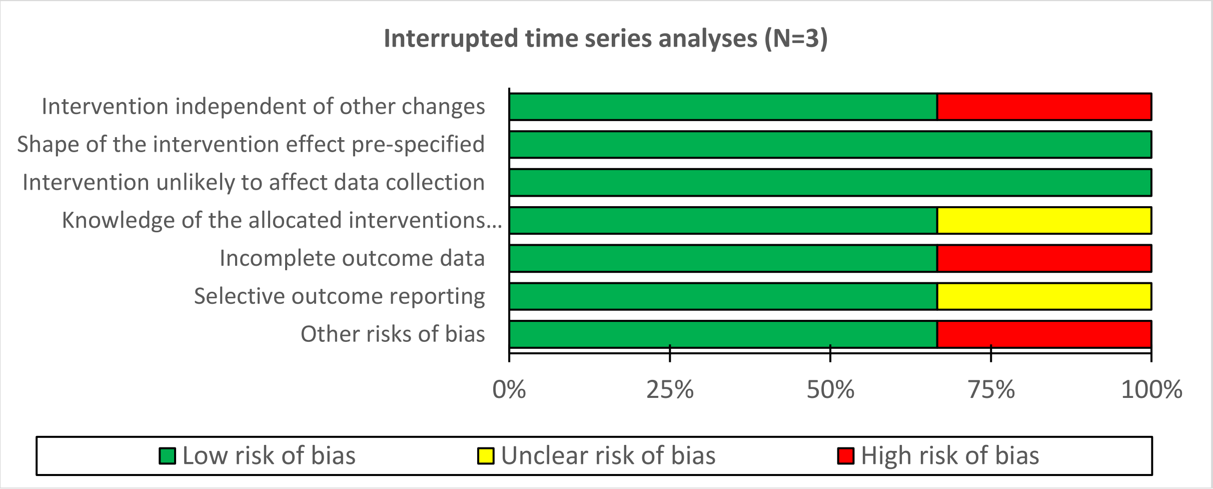

#### S8 Appendix: List of all endpoints for included studies

| Study ID | Outcome measure(s) |
|---|---|
| Aghlmandi 2023 | Primary:<br>Antibiotic prescription rate per 100 consultations in the second year of intervention.<br>Secondary:<br>1. Antibiotic prescription rate per 100 consultations in the first year of intervention and over 2 years while considering 2 repeated measurements, over the first and the second year of intervention.<br>2. Use of broad-spectrum antibiotics per 100 patient consultations.<br>3. All-cause hospitalizations and infection-related hospitalizations.<br>4. Antibiotic use in 3 specific patient age groups (≤5 years, 6-65 years, and >65 years). |
| Curtis 2021 | Primary:<br>Difference in the proportion of practices having their dashboard viewed during the 15 week intervention period.<br>Difference in the proportion of antibiotics prescribed which were broad-spectrum.<br>Secondary:<br>1. Difference in the mean dashboard views per practice during the 15 week intervention period.<br>2. Number of practices accessing at least one link provided in the intervention, as a proportion of all practices contacted.<br>3. Number of links accessed at least once as a proportion of all links delivered by each method of contact.<br>4. Proportion of emails opened.<br>5. Descriptive analysis of browsing sessions arising from each link accessed.<br>6. Estimate the overall effect of the intervention on the number of broad-spectrum antibiotics prescribed.<br>7. Rate of total antibiotic prescribing per adjusted population unit.<br>8. Mean number of daily doses per prescription for uncomplicated urinary tract infections.<br>9. Mean number of trimethoprim items prescribed as a percentage of all nitrofurantoin and trimethoprim items. |
| Davdison 2023 | Monthly prescription rates were calculated as the number of encounters with an antibiotic prescription ordered, compared to the total number of eligible encounters (i.e., visits with relevant ICD-10 codes). |
| deLusignan 2021 | Changes in prescribing (continued/stopped) for metformin or aspirin in the people inappropriately prescribed these medications at baseline. |
| Dutcher 2021 | Presence of an antibiotic prescription at each in-person visit that included 1 or more ICD10-CM codes for a respiratory tract infection. |
| Guthrie 2016 | Primary:<br>Proportion of patients included in one or more of the defined six individual secondary outcomes (denominator) who receive any high risk prescription (numerator).<br>Secondary:<br>1. Proportion of patients aged 75 years and over who receive a prescription for an oral antipsychotic.<br>2. Proportion of patients aged 65 years and over and currently treated with a diuretic and an ACEi/ARB who receive a prescription an NSAID.<br>3. Proportion of patients aged 75 years and over who receive a prescription for an oral NSAID without co-prescription of a gastroprotective drug.<br>4. Proportion of patients aged 65 years and over and currently treated with aspirin or clopidogrel who receive a prescription for an oral NSAID without co-prescription of a gastroprotective drug.<br>5. Proportion of patients currently treated with an oral anticoagulant who receive a prescription for an oral NSAID without co-prescription of a gastroprotective drug. |

| | |
|---|---|
| | 6. Proportion of patients currently treated with an oral anticoagulant who receive a prescription for aspirin or clopidogrel without co-prescription of a gastroprotective drug. |
| Hemkens 2017 | Primary:<br>Prescribed DDD (the assumed average maintenance dose per day for a drug used for its main indication in adults) of any type of antibiotics to any patient per 100 consultations in the first year.<br>Secondary:<br>1. Antibiotic prescriptions were further assessed in young children (0 to 5 years), older children and adolescents (6 to 18 years), younger adults (19 to 65 years), elderly (older than 65 years), and women or men.<br>2. Prescriptions for specific antibiotic types (e.g. tetracyclines, amphenicols, β-lactams/penicillins).<br>3. Prescribed DDD (the assumed average maintenance dose per day for a drug used for its main indication in adults) of any type of antibiotics to any patient per 100 consultations in the second year. |
| MacBride-Stewart 2022 | Primary:<br>The mean number of patients per practice with any of the five defined secondary outcome measures, listed below.<br>Secondary:<br>1. Aged 5–11 years with >12 SABA inhalers per annum and no inhaler containing single-agent or combination ICS (average daily exposure of <200 mcg beclometasone or equivalent).<br>2. Aged 12–34 years with >12 SABA inhalers per annum and no inhaler containing single-agent or combination ICS [or average daily exposure of <400 mcg beclometasone or equivalent]).<br>3. Aged ≥35 years with >12 SABA inhalers per annum and no long-acting muscarinic antagonist [LAMA] and no inhaler containing single-agent or combination ICS (average daily exposure of <400 mcg beclometasone or equivalent).<br>4. Aged <35 years with single-agent LABA (≥1 LABA inhaler and no single-agent ICS inhaler (single-agent ICS inhaler if average daily exposure was <400 mcg beclometasone or equivalent)).<br>5. Aged ≥35 years with single-agent LABA (≥1 LABA inhaler and no LAMA and no single-agent ICS inhaler (single-agent ICS inhaler if average daily exposure was <400 mcg beclometasone or equivalent)). |
| Peek 2020 | Primary:<br>Prevalence of exposure to any of the 10 secondary potentially hazardous prescribing indicators.<br>Prevalence of exposure to any of the two inadequate blood-test monitoring indicators.<br>Secondary:<br>1. Rates of ongoing (existent for 30 or more days) exposure to any potentially hazardous prescribing and any inadequate blood-test monitoring.<br>2. Rates of new (arisen within the previous 30 days) exposure to any potentially hazardous prescribing and any inadequate blood-test monitoring.<br>3. Prescription of an oral NSAID without co-prescription of a gastroprotective drug in a patient aged >65 years.<br>4. Prescription of an oral NSAID without co-prescription of a gastroprotective drug in a patient with a history of peptic ulcer disease.<br>5. Prescription of an antiplatelet drug without co-prescription of a gastroprotective drug in a patient with a history of peptic ulcer disease.<br>6. Prescription of warfarin or NOAC in combination with an oral NSAID.<br>7. Prescription of warfarin or NOAC in combination with an antiplatelet drug without co-prescription of a gastroprotective drug. |

| | |
|---|---|
| | 8. Prescription of aspirin in combination with another antiplatelet drug without co-prescription of a gastroprotective drug.<br>9. Prescription of a non-selective beta-blocker to a patient with asthma.<br>10. Prescription of a LABA to a patient with asthma who is not also prescribed an ICS.<br>11. Prescription of an oral NSAID to a patient with heart failure.<br>12. Prescription of an oral NSAID to a patient with chronic renal failure (eGFR < 45 ml/min/1.73 m2).<br>13. Prescription of methotrexate without both a recent full blood count and a liver function test in the last 3 months.<br>14. Prescription of amiodarone for at least 6 months without a thyroid function test within the last 6 months. |
| Soucy 2024 | Primary<br>1. Overall antibiotic prescribing rate (percentage of total visits with an antibiotic prescription).<br>2. Mean duration of therapy per antibiotic prescription.<br>3. Antibiotic prescribing rate for viral respiratory conditions (percentage of visits for viral respiratory conditions with an antibiotic prescription).<br>4. Antibiotic prescribing rate for acute sinusitis (percentage of visits for acute sinusitis with an antibiotic prescription). |
| Willis 2020,<br>Risk prescribing | Proportion of patients prescribed one or more of the defined nine individual secondary outcomes.<br>Secondary:<br>1. Prescription of an NSAID or low-dose aspirin in patients with a history of peptic ulceration without co-prescription of gastro-protection.<br>2. Prescription of an NSAID in patients aged 75 years or over without co-prescription of gastro-protection.<br>3. Prescription of an NSAID and aspirin in patients aged 65 years or over without co-prescription of gastro-protection.<br>4. Prescription of aspirin and clopidogrel in patients aged 65 years or over without co-prescription of gastro-protection.<br>5. Prescription of warfarin and an NSAID.<br>6. Prescription of warfarin and low-dose aspirin or clopidogrel without co-prescription of gastroprotection.<br>7. Prescription of an NSAID in patients with heart failure.<br>8. Prescription of an NSAID in patients prescribed both a diuretic and an ACEi/ARB).<br>9. Prescription of an NSAID in patients with chronic kidney disease (CKD). |
| Willis 2020,<br>Atrial fibrillation | Combined proportion of men with AF and a CHA2DS2-VASc score of 1 and women with a CHA2DS2-VASc score of 2 or above prescribed anticoagulation therapy. |

*Abbreviations: ICD-10, International Classification of Diseases; ICD-10-CM, International Classification of Diseases, Clinical Modification; ACEi, angiotensin-converting enzyme inhibitor; ARB, angiotensin II receptor blocker; NSAID, non-steroidal anti-inflammatory drug; DDD, defined daily dose; SABA, short-acting beta-agonist; ICS, inhaled corticosteroid; LAMA, long-acting muscarinic antagonist; LABA, long-acting beta-agonist; NOAC, non-vitamin K antagonist oral anticoagulants; eGFR, estimated glomerular filtration rate; CKD, chronic kidney disease; AF, atrial fibrillation; CHA2DS2-VASc, Congestive heart failure, Hypertension, Age ≥75 years, Diabetes mellitus, prior Stroke, Vascular disease, Age 65–74 years, and Sex category (female).*
